# Equal, equitable or exacerbating inequalities? Patterns and predictors of social prescribing referrals in 160,128 UK patients

**DOI:** 10.1101/2024.03.26.24304896

**Authors:** Feifei Bu, Daniel Hayes, Alexandra Burton, Daisy Fancourt

## Abstract

**Background:** Social prescribing (SP) is growing rapidly globally as a way to tackle social determinants of health. However, whom it is reaching and how effectively it is being implemented remains unclear.

**Aims:** To gain a comprehensive picture of SP in the UK, from referral routes, reasons, to contacts with link workers and prescribed interventions.

**Methods:** This study undertook the first analyses of a large database of administrative data from over 160,000 individuals referred to SP across the UK. Data were analysed using descriptive analyses and regression modelling, including logistic regression for binary outcomes and negative binomial regression for count variables.

**Results:** Mental health was the most common referral reason and mental health interventions the most common interventions prescribed. Between 72% and 85% of SP referrals were from medical routes (primary or secondary health care). While these referrals demonstrate equality in reaching across socio-demographic groups, individuals from more deprived areas, younger adults, men, and ethnic minority groups were reached more equitably via non-medical routes (e.g. self-referral, school, charity). Despite 90% of referrals leading to contact with a link worker, only 38% resulted in any intervention being received. A shortage of provision of community activities - especially ones relevant to mental health, practical support, and social relationships - was evident. There was also substantial heterogeneity in how SP is being implemented across UK nations.

**Conclusions:** Mental health is the leading reason for SP referrals, demonstrating its relevance to psychiatrists. But there are inequalities in referrals. Non-medical referral routes could play an important role in addressing inequality in accessing social prescribing, therefore should be prioritised. Additionally, more financial and infrastructural resource and strategic planning are needed to address low intervention rates. Further investment into large-scale data platforms and staff training are needed to continue monitoring the development and distribution of social prescribing.

## Introduction

Social determinants of health (SDOH), including income, education, employment, housing, childhood experience, social support, have long been demonstrated to play an important role in determining health outcomes.^1–3^ SDOH are estimated to account for 47% of health outcomes compared to just the 16% attributed to clinical care.^1^ Social prescribing (SP) is proposed as a way to address SDOH and improve health and wellbeing outcomes.^4^ Although SP has been advocated and practiced since 1980s, not until the last decade has it been widely recognised and implemented across the UK.^5^ SP involves linking patients with non-medical forms of support in their local communities, such as physical activity, social support, mental health support, financial advice, housing support, and so forth.^6^ The predominant SP model involves a healthcare professional referring patients on to a Link Worker, Community Connector, Community Navigator, or Health Trainer, who then works with the patient to develop a personalised care plan which connects them to community support and interventions.^4^

Evidence of mixed quality suggests that SP helps with a wide range of problems, including supporting mental health, wellbeing and health behaviours.^7–10^ Preliminary evidence also suggests that SP either reduces healthcare demand and costs or moves healthcare demand to primary care services rather than more expensive tertiary care, and demonstrates a favourable return on investment.^11^ Given its potential, NHS England has committed to tripling the number Link Workers by 2036/37.^12^ However, despite the promises of SP, national evaluations remain scant. Instead, evidence on SP is often limited to small scale, local evaluations and homogenous patient groups.^13–16^ Reviews into the use of SP suggest that referrals are not equitable, with those who are under 16, male, and those from non-white backgrounds being less likely to be offered SP.^14^ There are even concerns that SP is not effective for addressing social inequalities – it cannot address upstream determinants of inequalities and could even be provided disproportionately more to individuals who are less disadvantaged.^17–19^

To try and better understand the national picture of SP, studies have begun to explore patterns of SP in electronic patient health records. For example, the SP Observatory has analysed administrative data on SP referrals from over 1800 GP practices in England, showing a rapid increase in referrals since 2020, especially during the COVID-19 pandemic.^20^ Significant variation in rates of referrals across England has been identified, but with no substantial variation in referral rates by gender, ethnicity or neighbourhood deprivation.^20^ Whilst findings such as this are of great importance, there are limitations. First, SP is not limited to GP referrals, with referrals also made in secondary care,^21^ through voluntary and community sector organisations (VCSOs), via statutory services (e.g. education, social care), self-referral and more.^22^ Currently, the demographics of SP referrals via non-medical pathways are largely unknown. Moreover, given that certain demographics, particularly people from ethnic minority backgrounds, are less likely to report positive experiences accessing GP services ^23^ and young people report not wanting to access GP support for their mental health,^24^ it is important to establish if such groups are accessing SP via these alternative pathways. Second, there is only very limited information available in SNOMED codes used by GP practices, and these can include inconsistent coding, alongside poor recording of wider determinants of health leading to conclusions that they cannot be used to assess equity of referrals.^20,25^ Indeed, little is known about if SP referrals lead to any contact with link workers and actual interventions, why people are referred to SP, what interventions are prescribed, and what aspects lead to a successful, engaged SP referral. Third, national SP research has largely relied on data in England. How SP is implemented in other devolved countries in the UK, is also unknown.

The growth in cloud-based referral-management platforms that allow multi-stakeholder data input about SP pathways and integrate with primary care, secondary care and social care systems, is presenting new opportunities for understanding how SP is working and who it is reaching.^26^ However, data from these platforms have not been properly capitalised on. Therefore, the present study was initiated to address the knowledge gaps outlined above by using administrative data from one such SP platform: Access Elemental. We aimed to explore demographic characteristics of people who were referred to SP, referral reasons (including the importance of mental health within this), number of contacts with link workers, and interventions being provided through SP pathways. We also aimed to explore whether these rates differed across medical and non-medical referral pathways and whether there were any early indications of differences in individual country SP models. Overall, these analysed aimed to provide a more comprehensive understanding of SP roll-out across the UK, in particular for psychiatrists, with important implications for future policy, service, and workforce planning.

## Method

### Data

Data were administrative records obtained from Access Elemental,^27^ a digital SP platform used by health and social care professionals, community development workers and other service providers to keep track of SP activities and their impact from the point of referral. As a paid service, Elemental is the most widely adopted SP platform in the UK to date. It serves a population of over 20 million people across England, Scotland, Wales and Northern Ireland, and is used by over 37,740 health and care professionals and 4,405 social prescribers. To date, Elemental has recorded over 2.1 million contacts between patients and SP teams, including over 438,000 referrals to community-based programmes, services, and interventions. While Elemental does not comprise a random sample of sites or patients in the UK, it does have good geographical coverage in England (45% of Integrated Care Boards), Wales (57% of Welsh University Health Boards) and Northern Ireland (80% of Health and Social Care Trusts), alongside smaller representation in Scotland (13% of Scottish Health and Social Care Partnerships). Further, due to the existence of multiple IT systems in primary care in the UK, it is typical for research involving primary care to only involve a proportion of the whole population.^28^

Elemental collects detailed information tracking service users’ SP journeys, including referral source, referral reason, appointment with link worker, prescribed interventions, and so forth. They also collect core demographic information, such as gender, age, ethnicity (optional). Elemental data are available since January 2017, and this study used data up until November 2022. In this time, there were 201,037 SP cases from 169,818 unique individuals. 94.3% of individuals provided valid postcodes (N=160,128), allowing for data linkage with publicly available geographic information (see Figure S1). Figure S2 shows the distribution of SP service users across local authorities in the UK. Figure S3 shows the number of SP service users over time in the UK and across different countries. Most cases (88%) occurred between 2020-2022 in the UK, after the formal launch of the scheme as part of the NHS England Long-Term Plan.^29^ The research project was approved by the UCL Research Ethics Committee (23909/001).

### Measurement

Referrals sources were used as binary medical vs non-medical referrals. Medical referrals were mainly from GP practices, but also secondary care staff such as occupational therapists, physiotherapists, discharge teams and hospital based social workers. Non-medical referrals could come from educational establishments, local authorities, VSCOs, housing associations, self-referral, and so forth.

Referral reasons were coded in 600+ categories and we grouped them into seven domains: 1) mental health and wellbeing, 2) physical health and wellbeing, 3) social relationships (e.g. loneliness and social isolation), 4) lifestyle (e.g. exercise, diet, drinking, smoking), 5) employment, education and skills, 6) practical support (e.g. housing, finance, legal), 7) other reasons (see Table S1 in the Supplement). The grouping was done by two of the authors independently and any disagreement was discussed and resolved by consensus. These domains were non-mutually exclusive, allowing people to have more than one referral reasons.

Once people were referred to SP, they were connected to a SP link worker or a professional in a similar role (see Figure S4). We were interested in how many contacts with a link worker were made related to each SP case, referred as the number of contacts.

Contacts with link workers might or might not lead to individuals being connected with suitable community resources, referred as interventions in the present study. The intervention variable concerns if any prescription was issued by the link worker and what activities it entailed, without addressing service attendance or engagement. To be consistent with referral reasons, interventions (400+ categories) were also grouped into seven non-mutually exclusive domains (Table S1): 1) mental health and wellbeing, 2) physical health and wellbeing, 3) social relationships, 4) lifestyle, 5) employment, education and skills, 6) practical support, 7) other support.

Socio-demographic covariates included gender (female, male, other), age (0-17, 18-29, 30-39, 40-49, 50-59, 60-69, 70-79, 80+), urbanicity (urban, rural) and index of multiple deprivation (deciles: 1-most deprived to 10-least deprived). Both urbanicity and area of deprivation were derived via postcode linkage to urban rural classification and index of multiple deprivation at Lower Layer Super Output Area (LSOA) in each country. Although ethnicity was available, as an optional measure it had a high missing rate (>83%), which was excluded from most analysis.

### Statistical analysis

Descriptive analyses were undertaken to compare the socio-demographic profiles of people referred to SP services across UK countries, the percentages of different referral reasons, types of interventions received, and to what extent interventions matched to referral reasons. Regression analyses were conducted to assess variables that were associated with SP services stratified by country, excluding Northern Ireland due to a large proportion of missing data (82.9%) on referral route (Table S2). Referral route and interventions received were analysed using logistic regression models. The number of contacts with link workers was modelled using negative binomial regression to account for over-dispersion. All models controlled for socio-demographic covariates. A small percentage of people had more than one SP case (<14%), so the most recent record was used in regression analyses. Missing data were excluded in descriptive analyses but imputed in regression models by using multiple imputation with chained equations, including all covariates (m=30). All analyses were conducted in Stata V18.

## Results

### Demographic profiles

Among 160,128 unique individuals referred to SP, 141,011 people (88.1%) lived in England, 9,613 (6.0%) in Wales, 2,374 (1.5%) in Scotland and 7,130 (4.5%) in Northern Ireland. This proportion was largely similar to national statistics, with a slight overrepresentation of people from England (Table S3 in the Supplement). There was an overrepresentation of females being referred to SP in all countries, especially in Northern Ireland where 72.8% were females (Figure 1a, Table S4). Most service users (78.0%-87.2%) were aged 18 to 69 in each country (Figure 1b). However, Wales (5.2%) and Northern Ireland (11.5%) had a higher percentage of people aged under 18. Scotland had a higher percentage of people in their 60s (32.7%). Only 5.6% of SP service users lived in rural areas in England, much lower than the rural population estimate of 17.1% (Figure 1c). However, this percentage was substantially higher in Wales (24.1% vs 20% rural population), Scotland (27.2% vs 17% rural population) and Northern Ireland (35.5% vs 36% rural population). SP service users were more likely to live in deprived areas (Figure 1d). In the general population, roughly 10% of people living in each deprivation decile in each country. In England, 45.9% of people referred to SP lived in the top three most deprived areas, 45.8% in Wales and 63.8% in Northern Ireland. However, in Scotland, service users were more evenly distributed across deprivation deciles, except for the top two least deprived deciles.

**Figure 1.**
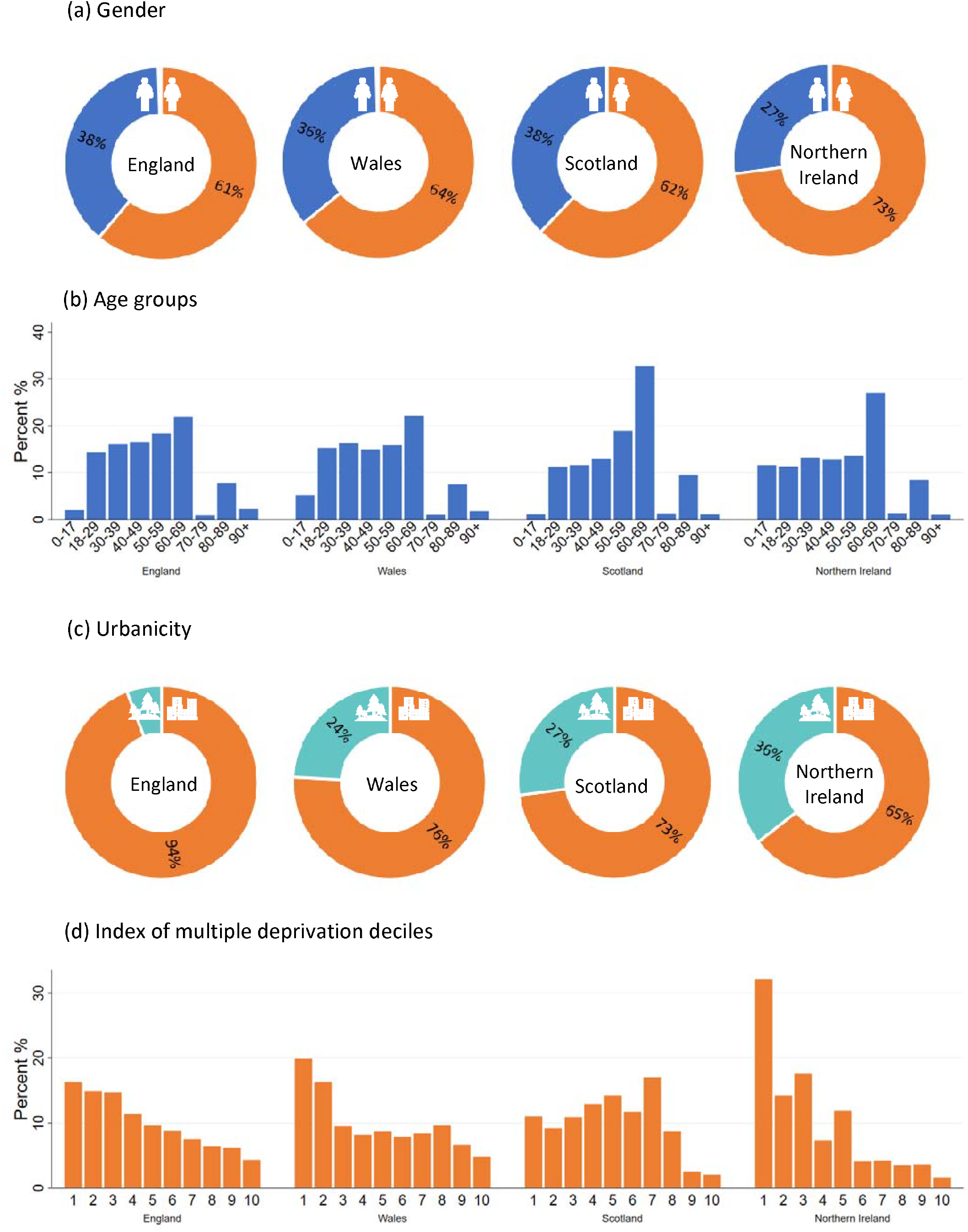
Socio-demographic characteristics of unique individuals by country

### Referral routes

Most SP cases were referred from medical routes in England (85.3%), Wales (72.3%) and Scotland (84.8%). The percentage was much lower in Northern Ireland (22.8%), possibly due to a high missing rate (82.9%, see Table S2). Table S5 in the Supplement compares socio-demographic profiles of people referred from medical and non-medical routes. Notably, SP service users referred via medical routes were more evenly distributed across ethnic groups, and deprivation deciles; whereas those from non-medical routes were more balanced across gender and urbanicity groups. All socio-demographic factors were related to referral routes, but estimated associations differed across countries (Figure 2 & Table S6). Key country differences were summarised in Table 1.

**Table 1.** Summary of key differences across countries from regression analyses.

|  | England | Wales | Scotland |
| --- | --- | --- | --- |
| Referral routes<br>(Figure 2, Table S6) | • Males were less likely to be referred via medical routes.<br>• People from older age groups (40+) had higher odds of being referred via medical routes (vs. aged 30-39).<br>• Urban residents had 3.24 times higher odds of being referred via medical routes.<br>• People from less deprived areas were more likely to be from medical routes. | • Males were less likely to be referred via medical routes.<br>• Children and young adults (0-29) were more likely to be referred via medical routes (vs. aged 30-39).<br>• Urban residents had 1.63 times higher odds of being from medical routes.<br>• People from less deprived areas were less likely to be from medical routes. | • No difference by gender.<br>• Children under 18 and older adults in their 60s were less likely to be referred via medical routes (vs. aged 30-39).<br>• No difference by urbanity.<br>• People from less deprived areas were less likely to be from medical routes. |
| Contacts with link workers<br>(Figure S7, Table S7) | • Males had fewer contacts.<br>• Urban residents had fewer contacts.<br>• Older adults aged 60+ had fewer contacts.<br>• People from less deprived areas had more contacts | • Males had fewer contacts.<br>• Urban residents had fewer contacts.<br>• Older adults aged 80+ had fewer contacts.<br>• People from less deprived areas had more contacts, but no difference between the most and least deprived deciles | • Males had more contacts.<br>• No difference by urbanicity.<br>• Older adults in their 60s and those aged 80+ had more contacts.<br>• People from less deprived areas had more contacts, but no difference between the most and least deprived deciles |
| Interventions<br>(Figure S8, Table S8) | • Those from medical routes were less likely to have an intervention.<br>• People with employment related reasons had 2.52 times higher odds of having an intervention; whereas those with lifestyle or practical reasons had lower odds.<br>• Male were less likely to have an intervention.<br>• Children under 18 were less likely to have an intervention. People aged 40-69 were more likely to have an intervention.<br>• Urban residents had 1.27 times higher odds of having an intervention.<br>• People from less deprived areas were less likely to have an intervention. | • Those from medical routes were more likely to have an intervention.<br>• People with employment and practical reasons had higher odds of having an intervention, lower odds for lifestyle reasons.<br>• No difference by gender.<br>• Children under 18 were more likely to have an intervention. No difference between those aged 40-69 and 30-39.<br>• Urban residents were less likely to have an intervention.<br>• People from less deprived areas were more likely to have an intervention, except for the top three least deprived deciles. | • Those from medical routes were less likely to have an intervention.<br>• People with lifestyle reasons had 2.03 times higher odds of having an intervention, but no difference by employment or practical reasons.<br>• No difference by gender.<br>• Children under 18 were more likely to have an intervention. No difference between those aged 40-69 and 30-39.<br>• Urban residents were less likely to have an intervention.<br>• Limited difference by area deprivation. |

**Figure 2.**
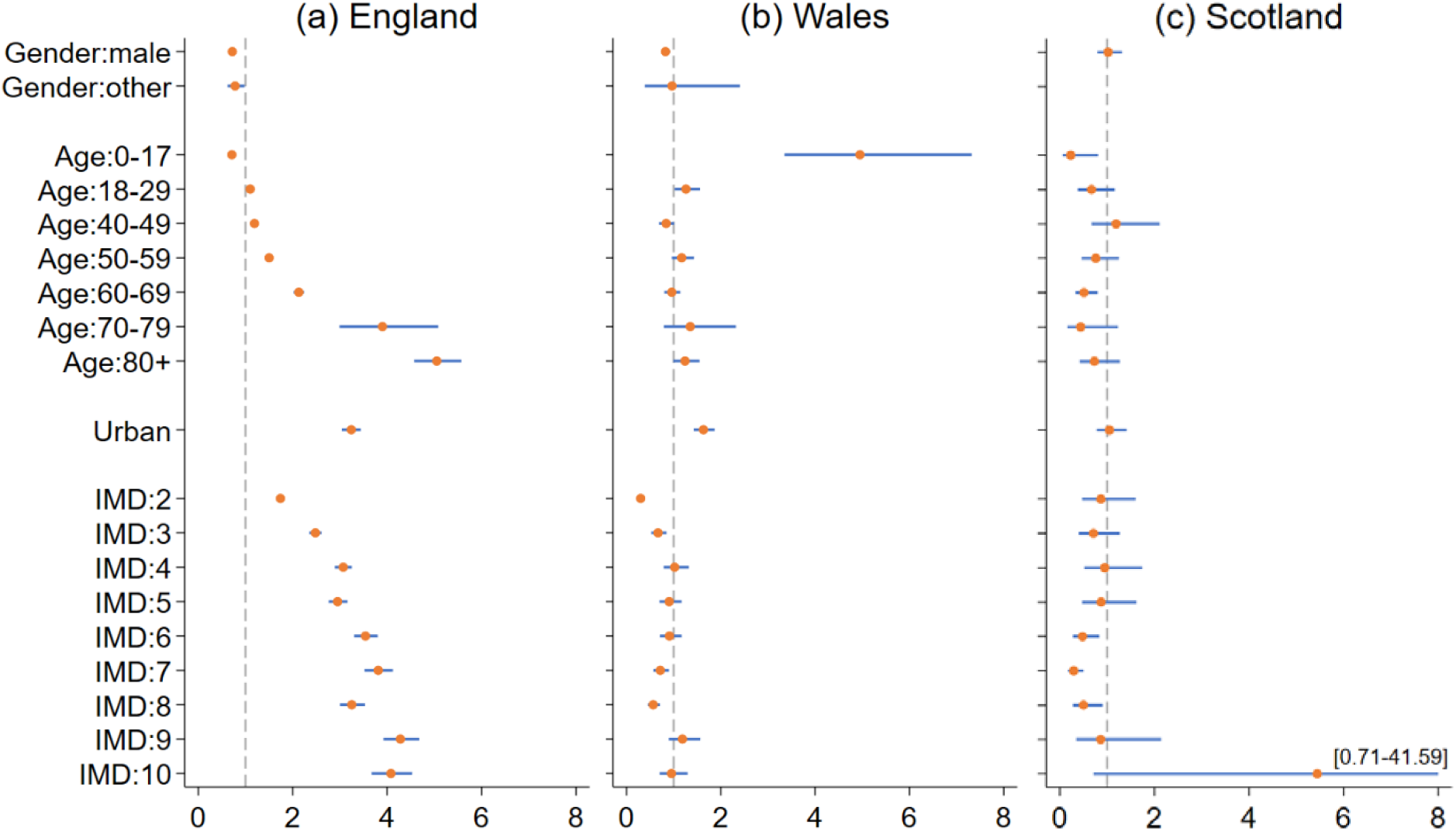
Odds ratios and 95% confidence intervals from the logistic regression model on being referred via medical routes by country

### Referral reasons

The most common referral reasons were related to mental health and wellbeing (33.5%), followed by practical support (26.1%) and social relationship (22.5%); whereas employment or education/skills (10.3%) and physical health (16.4%) related reasons were relatively less common (Figure 3a). Most SP cases (69.7%) reported reasons from a single domain only. About a third reported reasons from more than one domain. As shown in Figure 3b, 60.3% of cases referred for mental health reasons also had reasons from other domains, 52.3% practical support, 63.4% social support and 50.7% physical health. Figure S5 shows differences across countries. Physical health was among the most common reason in Wales, Scotland and Northern Ireland. Lifestyle related reasons were also common in Scotland and Northern Ireland.

**Figure 3.**
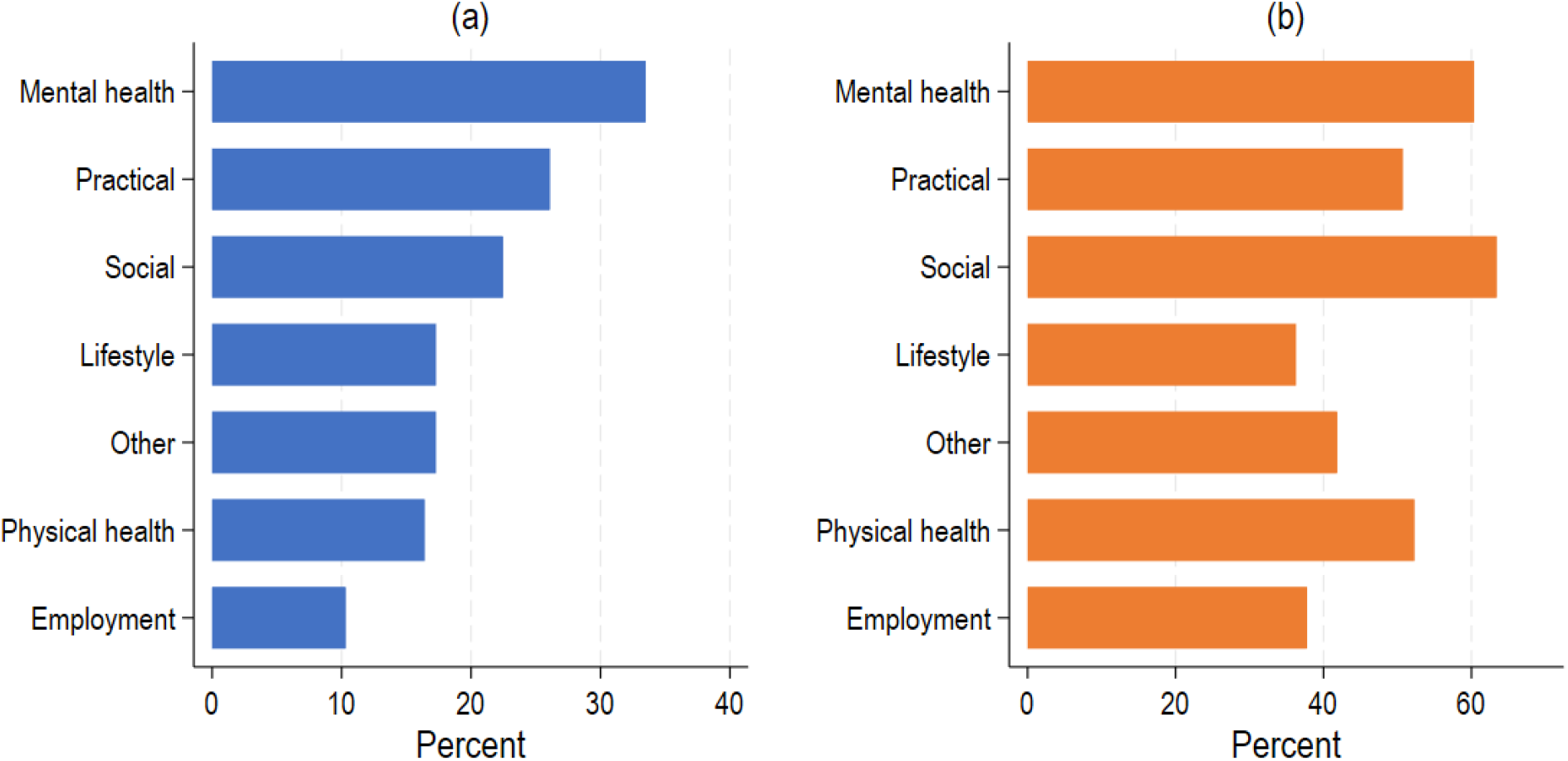
Referral reasons (a) percentage of cases with referral reasons from each domain (b) percentages of cases with reasons from one or more different domain for each domain

### Contacts with link workers

After being referred, each case would be assigned to a link worker who typically initiated contacts. Approximately 90% of SP cases had at least one contact with a link worker. Among cases without any recorded contact with a link worker, 53.1% were active cases who might yet to be contacted. On average, there were 4.2 contacts per case (SD=6.8, median=2). Among all the contacts, 11.5% were unsuccessful, 31.6% having reached clients with specific outcomes (e.g. appointment arranged, referral made, signposted, declined service), and 56.9% unclear (e.g. contact made, email/message left). Most of contacts (81.2%) were made via phone, video, email or letter, only 6.3% face-to-face, and another 12.6% unspecified. The contacts typically lasted for 0-15 minutes (59.4%), but some were longer (Figure S6). People referred from a medical route had a higher number of contacts across all countries (Figure S7 and Table S7). Gender, age, urbanity and area deprivation were also associated with the number of contacts, but some heterogeneities were founded across countries (Table 1).

### Interventions

Among 201,037 cases, 38.3% (n=76,992) had at least one intervention prescribed. Intervention prescription rates were 35.4% in England, 33.0% in Wales, but much higher in Scotland (51.9%) and Northern Ireland (74.9%). Referral routes, number of contacts, referral reasons, age, urbanity were associated with the odds of having an intervention prescribed (Figure S8 and Table S8). But there were notably differences across countries (Table 1). For example, urban residents were more likely to have an intervention in England, but less likely in Wales and Scotland than rural residents.

For SP cases with at least one intervention (n=76,971), there were 165,595 prescribed interventions (2.2 per case). Most of these interventions (91.7%) were recorded as free to access. Only 6.3% had out-of-pocket costs and another 1.9% were subsidised (Table S9). England and Wales were similar in that over 95% interventions were free. But only 50.3% interventions were free in Scotland and 78.7% in Northern Ireland (Table S9).

Most prescribed interventions (82.9%) had information on service domains, with 62.9% of these interventions falling under one service domain and 37.1% with more than one domain. As shown in Figure 4a, around 43.8% SP cases had an intervention related to mental health and wellbeing, followed by social relationships (36.3%), lifestyle (28.7%), employment, education and skills (25.4%), practical support (24.6%), physical health and wellbeing (19.0%) and other support (12.7%). Linking back to referral reasons, Figure 4b shows the percentage of cases with an intervention prescribed by referral reasons in the UK. 69.3% of cases who were referred due to mental health reasons had no intervention, 67.6% for physical health, 71.0% for social relationship, 65.9% for lifestyle, 70.5% for practical reasons, 72.8% for other reasons. The percentage of cases with no intervention was much lower for those referred for employment related reasons (37.1%). Among cases who did have an intervention, those who were referred for lifestyle related reasons were more likely to have an intervention matched to these reasons (81.7%), followed by employment (76.1%), mental health (70.3%), practical support (60.5%) and social relationships (60.1%) reasons. Cases for physical health (32.7%) or other reasons (14.9%) were less likely to have a matched intervention, but the patterns differed across countries (Figure S9). For instance, 35.9% of cases who were referred due to lifestyle reasons had no intervention in Scotland, compared to 20.8% in Northern Ireland. Moreover, only 18.6% of cases with social reasons had no intervention in Northern Ireland.

**Figure 4.**
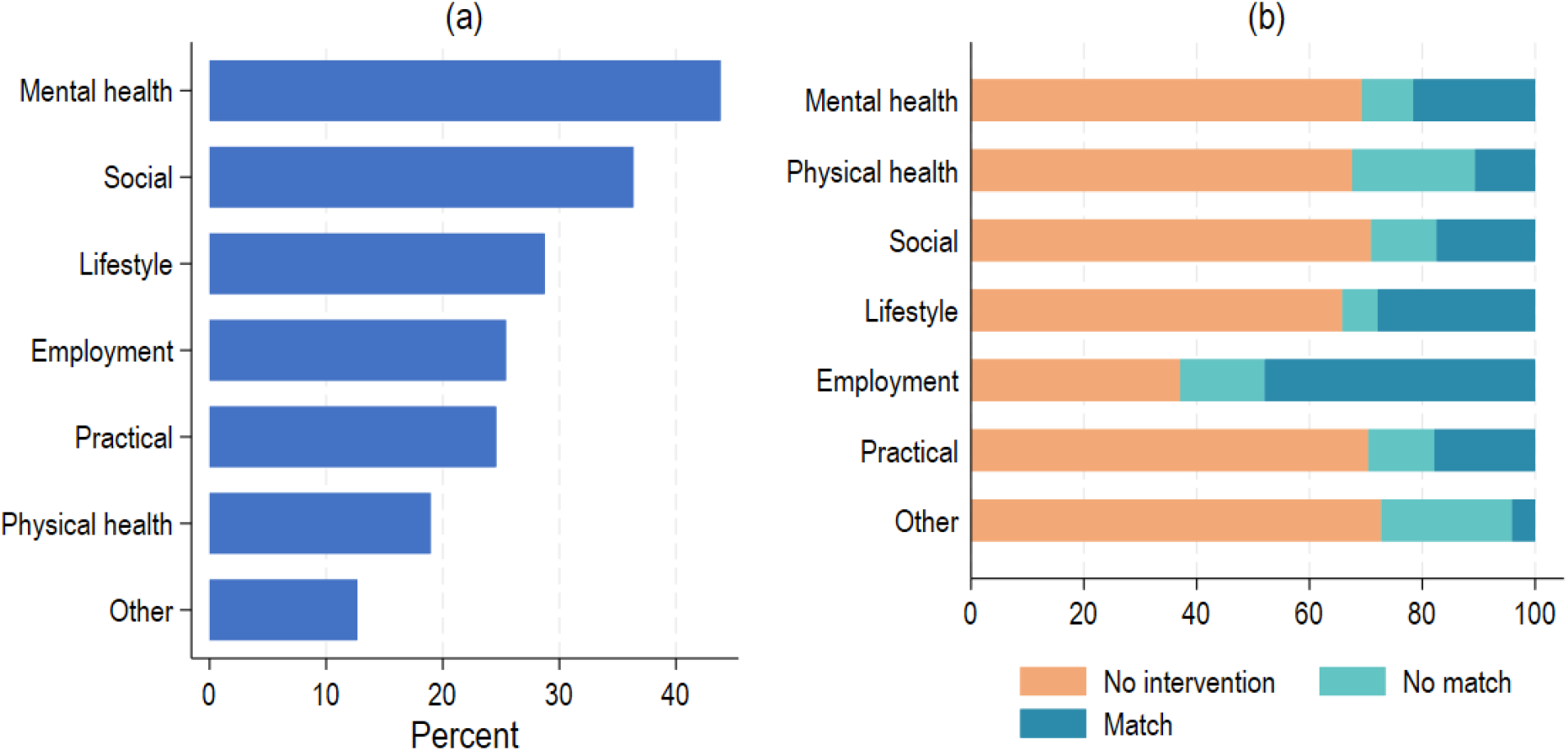
Intervention domains (a) among cases with an intervention, percentage of cases belong to each domain (b) percentages of cases receiving an intervention matched to referral reason in each referral reason domain

## Discussion

This study is the first UK-wide study on SP referrals in the UK, with fine details on service users’ socio-demographic profiles, referral routes, reasons, contacts with link workers and actual interventions and their costs. While relying on data from just one data management system, it provides importance evidence to triangulate with previous research using GP records and smaller-scale evaluations by providing (i) simultaneous data on both medical and non-medical referral sources, allowing the first direct comparison of population characteristics, (ii) detail not just on whether a referral was made but the process by which the referral pathway was carried out, and (iii) a large sample size with data over several years. Most notably, it highlights that mental health is the leading cause of referrals across the UK. It also systematically investigates the large-scale differences between medical and non-medical SP pathways across the UK and provides early data on differences across the devolved countries in the UK. Understanding of the status-quo of SP in the UK is crucial for identifying its future directions and supporting the roll-out of SP as a public health intervention around the world.

As expected, the medical pathway is the dominant referral route in the UK. The seemingly exception of Northern Ireland is likely to be driven by missing data. The medical pathway has its merits. Everyone in the UK is entitled to free healthcare, and most people (if not all) are registered with a GP. This may explain why SP service users were more evenly distributed across deprivation deciles and there was a higher percentage of people from ethnic minority background among people from the medical route compared to non-medical routes. This is consistent with previous findings of no ethnic or area deprivation difference in SP when focusing on primary care data.^20^ In this vein, the medical pathway prevails in promoting equality. However, when it comes to actual service utilisation, there are notable socio-demographic differences. For example, there is less use of primary healthcare services among young people and men,^30,31^ which may limit their chances of being referred to SP through primary care. In addition to equality, we need to consider equity. People from deprived areas generally have higher risks of health problems and more complex needs, potentially requiring extra support and resources to achieve an equal outcome.^32,33^ Our findings suggest that the non-medical pathway complements the medical pathway by reaching the underrepresented in healthcare and those who are in greater need of support (e.g. those from more deprived areas). The two pathways should work hand in hand to maximise the overall impact and efficiency of SP.

Referral, regardless of route, is only an early step in the lifecycle of an SP case. A key question is whether a referral leads to any intervention. Our analyses revealed that 9 in 10 referred SP cases had at least one contact with a link worker, showing a successful initial transition. However, only 38.3% of cases were subsequently referred to an intervention. What is unclear is whether this low level of intervention engagement was voluntary (i.e. patients finding sessions of talking and a supportive relationship sufficient for their needs) or due to other issues such as a Link Worker not being able to reach the client, referral being declined, or a lack of appropriate resources or suitable interventions in the local community (especially considering that over 90% of prescribed interventions are free of charge so depend on such provision being available). However, several factors suggest that it is not entirely voluntary. First, referral to an intervention is not even. People living in urban areas are more likely to be referred to an intervention than rural residents, suggesting that a lack of provision may be an issue. Additionally, although mental health, practical support and social relationship are the most common referral reasons, the percentage of people having a matched intervention for each of these reasons is only around 20%, compared to 47.9% for employment – provision of free employment support which has long been a priority in the UK, suggesting that when free interventions are readily available, uptake is higher.^34^ Therefore, there is urgent work that is needed either building further community infrastructure in domains outside employment to enable free services to be available, or providing further ringfenced funding for link workers to utilise in under-resourced intervention domains. Additionally, more behavioural research is recommended to identify what the barriers to accessing interventions are and how they can be addressed.

Another finding that stands out is the heterogeneity of SP across countries in the UK. Socio-demographic profiles of SP service users across countries. For example, in Wales and Northern Ireland, there are higher percentages of children and young people under 18, as well as people from the most deprived decile being referred to SP. This could reflect different approaches being adopted with respect to equality and equity in different countries, which is in line with our finding of differential associations of socio-demographic factors with referral routes, contacts with link workers and prescribed interventions across countries. Differences in service user compositions could also, to some extent, explain the country differences in the distribution of referral reasons. For example, over 50% of SP cases are referred due to practical reasons in Wales, and lifestyle is the most common referral reason in Northern Ireland (one of the least common reasons in England and Wales). Intervention rates are notably higher in Scotland and Northern Ireland, which could be related to reasons of referral and the availability of required services. Notably, England and Wales have a much lower percentage of interventions that are costed or subsidised than Scotland and Northern Ireland, providing further evidence that relying on free services due to a lack of funding could be a major obstacle to an efficient and effective service.

However, with regards to limitations, the country differences should be interpreted with caution as we are unable to assess the representativeness of the data. This is particularly concerning for Wales, Scotland and Northern Ireland where the sample sizes are small. Although Access Elemental is the largest digital platform for SP across the UK, we cannot rule out the possibilities that the country differences might be driven by different clienteles of the Access Elemental in these countries compared to England. Indeed, despite being used across a diverse range of places in the UK, Elemental has a low coverage in Scotland (18%) and is not used in some areas e.g. Greater Glasgow and Clyde in Scotland, which have high levels of deprivation, accounting for the much lower representation we had in our dataset of people from the lowest two deciles of deprivation in Scotland. Given that all primary care data in the UK is non-representative given the multiple different data platforms in use both amongst GPs and for more specialist purposes like SP, this challenge is not easily solved, but further work cross-validating the findings presented here is encouraged if we are to fully understand cross-country differences in SP. In these analyses, although ethnicity is recorded in the data, the fact that it is ‘optional’ for data input means it is missing for most people, so we were unable to examine any SP differences related to ethnicity.

Additionally, there is limited information on people’s medical conditions which may influence the SP services that they receive. Continued efforts are needed to improve data quality and accessibility for a better understanding of SP implementation and its impacts. Our study has the strength of using data from the most widely adopted SP platform in the UK to date, serving a large and diverse population with detailed information about diverse referral routes. However, due to its nature, we were unable to compare our sample to those who have never been considered for SP. Development of further large-scale rigorous evaluations of SP as a service are needed.

Overall, this analysis shows that SP needs to be understood more by psychiatrists given the central role of mental health. It has previously been argued that SP is aligned with the principles of modern psychiatric care, emphasising what matters to the individual, prioritising belonging and community inclusion, supporting emotion processing and regulation, and facilitating behavioural adaptations.^35^ SP is also timely, given existing challenges in psychiatry such as medication adherence, availability of traditional psychiatric services, and shifts in thinking about the concept of recovery.^35^ Psychiatrists are encouraged to consider the suitability of social prescribing as an additional clinical service they can refer or signpost their patients to alongside traditional psychiatric services. This can be carried out through existing Link Worker models using Link Workers available within primary care. However, novel programmes embedding SP into psychiatric services such as in mental health service waiting lists in the UK and as part of shared decision making in psychiatric hospitals in Australia to support discharge plans are underway, and further such work is also encouraged.^36^ Nonetheless, although the medical pathway in SP is dominant across the UK, it has a number of challenges associated, including that it does not manage to reach certain demographic groups such as young people, men, and people from more deprived areas as effectively. Consequently, there is a clear need for non-medical referral routes to be prioritised in the policy and delivery planning and funding of future SP in the UK. Psychiatrists also have a role to play in supporting the development of non-clinical referral pathways for mental health, such as peer support SP pathways,^37^ to ensure that they are developed in a way that is safe, feasible, and appropriate to the mental health needs of those who will be using them.

Although SP is an individual-focused intervention that does not address the structural factors that may have given rise to those inequalities in the first place, results from non-medical referrals suggest SP could provide additional support to individuals experiencing inequalities.^38,39^ Additionally, despite a high number of SP referrals taking place, only a third are leading to an intervention after link worker consultations, with rates of interventions being particularly low on medical referral pathways and certain types of interventions (such as those for physical health) being very limited. Challenges in funding for community resources may be key here, with most financial resource for SP currently going to the pathway and affiliated staff rather than community resources. Additional financial resource for certain intervention domains for which there is not suitable free provision could help to readdress this balance. Third, given our findings of heterogeneity in how SP is being delivered, for whom, across England and devolved nations, specific strategies for the development of SP that meet the differing health and social needs in the different countries are recommended moving forwards. Finally, in order to be able to continue monitoring the development and distribution of SP in the UK, further investment into large-scale data platforms such as Access Elemental and staff training in good quality data capture is vital.

## Data Availability

Data are restricted access, currently only available through applications to Access Elemental

## Acknowledgments

We thank Access Elemental and all data holders for granting us the data access.

## Author contributors

DF and FB conceptualised the study and acquired the data. FB conducted the data analysis and designed the data visualisation. All authors (FB, DH, AB and DF) contributed to the interpretation of results and writing of the article, and had final responsibility for the decision to submit for publication. DF and FB directly accessed and verified underlying data.

## Data sharing

Due to the sensitive nature of the data, the research data can not be shared publicly. Data access can be requested from the Access Elemental subject to ethical restrictions.

