## Supplementary for "Equal, equitable or exacerbating inequalities? Patterns and predictors of social prescribing referrals in 160,128 UK patients"

Table S1. Examples of referral reason and intervention domains

|  | Referral reason | Intervention |
| --- | --- | --- |
| Mental health & wellbeing | Mental health, mental health issues, mental wellbeing, depression/anxiety , low self-esteem/confidence, recovery college, emotional wellbeing, bereavement , low mood/mild anxiety, self-harm, anti-social behaviour, stress management … | Mental health, recovery college, life rooms learning, mental wellbeing, Talking therapies/counselling, suicide prevention, bereavement, kind mind community, emotional support … |
| Physical health & wellbeing | COVID-19, long term condition management , physical wellbeing, fall prevention, dealing with chronic illness  pre-diabetic, general wellbeing , physical health, pain management, chronic pain, health & wellbeing coach, disability support … | Specific illness support, clinical support, physical health and wellbeing, health & wellbeing, dementia, disability support, COVID-19, cancer, pain management, general health, autism, population health outcomes … |
| Social relationships | Social isolation, social needs, loneliness and isolation, social interactions, social support , improve social contact, social and community, relationship breakdown, personal and social wellbeing, healthful network, relationship issues … | Social support, community activities-general, befriending & social isolation, social interactions, social & community, loneliness, social inclusion, support groups, reduce isolation … |
| Lifestyle | Physical inactivity, weight management/eating well, weight reduction, exercise referral , become more active, lifestyle-related problems, quit smoking, substance misuse, alcohol/drug problems … | Physical exercise, diet & nutrition, alcohol & substance misuse support, smoking support, lifestyle support, gym sessions, weight management … |
| Employment, education & skills | Skills training/course, personal development, employment and training, employment support, employability & preparation for work, volunteering, CV job application and interview preparation, motivation for learning, digital support, adult education … | Working well, employment support, employment & skills, learning and development, employment and training, adult education, self-employment, volunteering, education support, training courses … |
| Practical support | Housing, housing information, financial advice, benefits advice/support, benefits/money, help with basic daily needs, practical support, food support, food bank debt advice, homelessness, welfare support, access to transport, help with groceries, energy support, utility bills support, legal advice, carer, cares support … | Help with independent living, financial and benefits support, benefits/money, housing, housing support/advice, healthy homes healthy people, income maximisation, energy, financial shield support, information and advice, food support, food bank, social services, legal advice … |
| Other | Other, counselling, domestic violence, abuse, family support, safety, community support, refugee and asylum seeker advice, immigration support, children & young people services, advocacy, ex-offenders … | Social prescription, family support, advice services, children & young people, older people, signposting, helpline, family services, refugee/asylum seeker, advocacy services … |

Table S2. Social prescribing measures and missing data in the UK and by country

|  |  | UK  (N=201,037) | England  (N=167,690) | Wales  (N=11,567) | Scotland  (N=2,724) | Northern Ireland  (N=8,326) |
| --- | --- | --- | --- | --- | --- | --- |
| Referral route | Medical | 82.9% | 85.3% | 72.3% | 84.8% | 22.8% |
|  | Non-medical | 17.1% | 14.7% | 27.7% | 15.2% | 77.2% |
|  | Missing of total | 18.4% | 13.2% | 21.1% | 19.1% | 82.9% |
| Referral reasons | Mental health & wellbeing | 33.5% | 35.1% | 21.3% | 43.0% | 24.0% |
|  | Physical health & wellbeing | 16.4% | 15.1% | 28.9% | 35.5% | 23.3% |
|  | Social relationships | 22.5% | 23.0% | 18.4% | 24.0% | 19.4% |
|  | Life style | 17.3% | 17.2% | 3.7% | 33.0% | 27.1% |
|  | Employment, education & skills | 10.3% | 10.4% | 1.9% | 7.8% | 9.5% |
|  | Practical support | 26.1% | 26.0% | 50.9% | 14.6% | 7.2% |
|  | Other reasons | 17.3% | 18.2% | 12.2% | 13.4% | 10.5% |
|  | Missing of total | 2.3% | 1.7% | 2.4% | 4.2% | 10.7% |
| Number of contacts | Mean (SD) | 4.1 (6.0) | 4.3 (6.2) | 3.4 (4.5) | 4.4 (6.0) | 2.4 (4.6) |
|  | Missing of total | 0.0% | 0.0% | 0.0% | 0.0% | 0.0% |
| Interventions | Yes | 38.3% | 35.4% | 33.0% | 51.9% | 74.9% |
|  | No | 61.7% | 64.7% | 67.0% | 48.1% | 25.1% |
|  | Missing of total | 0.0% | 0.0% | 0.0% | 0.0% | 0.0% |

Table S3. Social prescribing service users in each country

| Country | Elemental data | ONS population estimates (Mid 2021)^†^ |
| --- | --- | --- |
| England | 88.1% | 84.3% |
| Wales | 6.0% | 4.6% |
| Scotland | 1.5% | 2.8% |
| Northern Ireland | 4.5% | 8.2% |

Note: ^†^ ONS https://www.ons.gov.uk/peoplepopulationandcommunity/populationandmigration/populationestimates/bulletins/annualmidyearpopulationestimates/mid2021

Table S4. Socio-demographic characteristics of unique individuals in the UK and by country

|  |  | UK  (n=160,128) | England  (n=141,011) | Wales  (n=9,613) | Scotland  (n=2,374) | Northern Ireland  (n=7,130) |
| --- | --- | --- | --- | --- | --- | --- |
| Gender | Male | 37.6% | 38.4% | 35.5% | 37.7% | 27.0% |
|  | Female | 61.9% | 61.1% | 64.1% | 62.1% | 72.8% |
|  | Other | 0.5% | 0.5% | 0.4% | 0.2% | 0.2% |
|  | Missing of total | 2.0% | 2.2% | 0.8% | 1.6% | 0.0% |
| Ethnicity | White | 75.4% | 73.3% | 100.0% | 93.7% | 98.9% |
|  | Ethnic minority | 24.6% | 26.7% | 0.0% | 6.3% | 1.1% |
|  | Missing of total | 85.1% | 83.8% | 97.6% | 86.7% | 87.4% |
| Age^‡^ | 0-17 | 3.0% | 2.0% | 5.2% | 1.1% | 11.5% |
|  | 18-29 | 14.2% | 14.3% | 15.2% | 11.2% | 11.3% |
|  | 30-39 | 15.9% | 16.1% | 16.3% | 11.5% | 13.2% |
|  | 40-49 | 16.2% | 16.5% | 14.9% | 12.9% | 12.8% |
|  | 50-59 | 17.8% | 18.4% | 15.9% | 18.9% | 13.6% |
|  | 60-69 | 22.2% | 21.9% | 22.1% | 32.7% | 27.0% |
|  | 70-79 | 0.9% | 0.9% | 1.0% | 1.2% | 1.3% |
|  | 80+ | 9.7% | 10.1% | 9.3% | 10.6% | 9.4% |
|  | Missing of total | 0.6% | 0.1% | 1.6% | 0.0% | 3.1% |
| Urbanicity^†^ | Urban | 91.6% | 94.3% | 75.9% | 72.8% | 64.5% |
|  | Rural | 8.4% | 5.7% | 24.1% | 27.2% | 35.5% |
|  | Missing of total | 5.7% | 0.0% | 0.0% | 0.9% | 0.0% |
| Area  deprivation^†^ | 1 (most deprived) | 17.2% | 16.3% | 19.9% | 11.0% | 32.1% |
|  | 2 | 14.8% | 14.9% | 16.3% | 9.2% | 14.2% |
|  | 3 | 14.5% | 14.7% | 9.5% | 10.9% | 17.6% |
|  | 4 | 11.1% | 11.4% | 8.2% | 12.9% | 7.3% |
|  | 5 | 9.7% | 9.6% | 8.7% | 14.2% | 11.9% |
|  | 6 | 8.6% | 8.8% | 7.9% | 11.7% | 4.1% |
|  | 7 | 7.5% | 7.5% | 8.4% | 17.0% | 4.2% |
|  | 8 | 6.5% | 6.4% | 9.6% | 8.7% | 3.5% |
|  | 9 | 6.1% | 6.2% | 6.6% | 2.5% | 3.6% |
|  | 10 (least deprived) | 4.1% | 4.3% | 4.8% | 2.0% | 1.6% |
|  | Missing of total | 5.7% | 0.0% | 0.0% | 0.9% | 0.0% |

Notes: † Urbanicity classification and index of multiple deprivation measures developed separately in each country, which are not necessarily comparable across countries. ‡ Age is taken as the most recent case for people with more than one record.

Table S5. Socio-demographic characteristics of unique individuals by referral routes (excluding Northern Ireland)

|  |  | Medical  (n=116,040) | Non-medical  (n=24,156) |
| --- | --- | --- | --- |
| Gender | Male | 36.8% | 44.8% |
|  | Female | 62.8% | 54.7% |
|  | Other | 0.4% | 0.5% |
|  | Missing of total | 1.0% | 1.1% |
| Ethnicity | White | 72.3% | 83.5% |
|  | Ethnic minority | 27.7% | 16.5% |
|  | Missing of total | 82.7% | 87.4% |
| Age^‡^ | 0-17 | 2.0% | 3.4% |
|  | 18-29 | 13.4% | 18.4% |
|  | 30-39 | 15.0% | 21.8% |
|  | 40-49 | 15.7% | 19.4% |
|  | 50-59 | 18.1% | 17.2% |
|  | 60-69 | 23.2% | 16.0% |
|  | 70-79 | 1.1% | 0.4% |
|  | 80+ | 11.6% | 3.4% |
|  | Missing of total | 0.6% | 0.3% |
| Urbanicity^†^ | Urban | 94.1% | 88.7% |
|  | Rural | 5.9% | 11.3% |
|  | Missing of total | 2.6% | 10.1% |
| Area  deprivation^†^ | 1 (most deprived) | 12.7% | 27.5% |
|  | 2 | 14.3% | 19.3% |
|  | 3 | 15.3% | 13.6% |
|  | 4 | 11.8% | 8.9% |
|  | 5 | 9.8% | 7.6% |
|  | 6 | 9.4% | 6.5% |
|  | 7 | 8.1% | 5.5% |
|  | 8 | 6.9% | 5.3% |
|  | 9 | 7.0% | 3.5% |
|  | 10 (least deprived) | 4.8% | 2.3% |
|  | Missing of total | 2.6% | 10.1% |

Notes: † Urbanicity classification and index of multiple deprivation measures developed separately in each country, which are not necessarily comparable across countries. ‡ Age is taken as the most recent case for people with more than one record.

Tables S6 Estimated odds ratios (ORs) and 95% Confidence intervals (CIs) from logistic regression models on medical referral route (vs other) for England, Wales and Scotland

|  | England  (n=141,011)  MI=30 | | Wales  (n=9,613)  MI=30 | | Scotland^†^  (n=2,370)  MI=30 | |
| --- | --- | --- | --- | --- | --- | --- |
|  | OR | 95% CI | OR | 95% CI | OR | 95% CI |
| Male (vs female) | **0.72** | **[0.69-0.74]** | **0.83** | **[0.74-0.92]** | 1.02 | [0.79-1.32] |
| Other (vs female) | **0.78** | **[0.62-0.98]** | 0.97 | [0.39-2.40] | -- | -- |
| Age: 0-17 (vs 30-39) | **0.71** | **[0.65-0.79]** | **4.95** | **[3.35-7.32]** | **0.23** | **[0.06-0.81]** |
| 18-29 (vs 30-39) | **1.10** | **[1.04-1.16]** | **1.26** | **[1.02-1.56]** | 0.67 | [0.38-1.16] |
| 40-49 (vs 30-39) | **1.19** | **[1.13-1.25]** | 0.84 | [0.69-1.01] | 1.19 | [0.67-2.11] |
| 50-59 (vs 30-39) | **1.50** | **[1.42-1.58]** | 1.17 | [0.96-1.43] | 0.76 | [0.46-1.25] |
| 60-69 (vs 30-39) | **2.13** | **[2.02-2.24]** | 0.96 | [0.80-1.14] | **0.51** | **[0.33-0.80]** |
| 70-79 (vs 30-39) | **3.90** | **[2.99-5.08]** | 1.35 | [0.79-2.32] | 0.44 | [0.16-1.23] |
| 80+ (vs 30-39) | **5.05** | **[4.57-5.57]** | 1.24 | [0.99-1.55] | 0.73 | [0.42-1.27] |
| Urban (vs rural) | **3.24** | **[3.04-3.44]** | **1.63** | **[1.42-1.87]** | 1.05 | [0.78-1.41] |
| Deprivation: 2 (vs 1) | **1.74** | **[1.66-1.83]** | **0.30** | **[0.25-0.37]** | 0.87 | [0.47-1.60] |
| 3 (vs 1) | **2.48** | **[2.35-2.61]** | **0.67** | **[0.52-0.85]** | 0.71 | [0.40-1.27] |
| 4 (vs 1) | **3.07** | **[2.89-3.25]** | 1.02 | [0.79-1.32] | 0.95 | [0.52-1.74] |
| 5 (vs 1) | **2.95** | **[2.76-3.16]** | 0.90 | [0.70-1.17] | 0.87 | [0.47-1.62] |
| 6 (vs 1) | **3.54** | **[3.30-3.80]** | 0.91 | [0.71-1.17] | **0.48** | **[0.28-0.84]** |
| 7 (vs 1) | **3.81** | **[3.52-4.12]** | **0.72** | **[0.57-0.90]** | **0.29** | **[0.17-0.50]** |
| 8 (vs 1) | **3.25** | **[3.00-3.53]** | **0.57** | **[0.45-0.71]** | **0.50** | **[0.28-0.91]** |
| 9 (vs 1) | **4.28** | **[3.92-4.68]** | 1.18 | [0.90-1.57] | 0.86 | [0.35-2.14] |
| 10 (vs 1) | **4.08** | **[3.67-4.53]** | 0.95 | [0.70-1.30] | 5.44 | [0.71-41.59] |

Notes: Ethnicity is excluded from the analysis due to large percentage of missing data. † The ‘other’ gender category is excluded for Scotland due to its small number.

Tables S7 Estimated incident rate ratios (IRRs) and 95% Confidence intervals (CIs) from negative binomial regression models on case visits by country

|  | England  (n=141,011)  MI=30 | | Wales  (n=9,613)  MI=30 | | Scotland^†^  (n=2,370)  MI=30 | |
| --- | --- | --- | --- | --- | --- | --- |
|  | IRR | 95% CI | IRR | 95% CI | IRR | 95% CI |
| Medical route (vs other) | **2.29** | **[2.25-2.33]** | **1.10** | **[1.04-1.16]** | **1.48** | **[1.30-1.68]** |
| Male (vs female) | **0.98** | **[0.97-0.99]** | **0.87** | **[0.83-0.91]** | **1.15** | **[1.06-1.26]** |
| Other (vs female) | 1.07 | [0.99-1.16] | 0.90 | [0.65-1.24] | -- | -- |
| Age: 0-17 (vs 30-39) | **0.62** | **[0.60-0.65]** | **0.83** | **[0.74-0.92]** | **0.28** | **[0.17-0.48]** |
| 18-29 (vs 30-39) | **0.97** | **[0.95-0.99]** | 0.99 | [0.92-1.07] | 0.93 | [0.78-1.11] |
| 40-49 (vs 30-39) | 1.00 | [0.98-1.02] | 0.96 | [0.89-1.03] | 1.08 | [0.91-1.28] |
| 50-59 (vs 30-39) | **1.02** | **[1.00-1.04]** | 1.03 | [0.96-1.11] | 1.04 | [0.89-1.21] |
| 60-69 (vs 30-39) | **0.96** | **[0.94-0.98]** | 0.94 | [0.88-1.01] | **1.36** | **[1.18-1.56]** |
| 70-79 (vs 30-39) | **0.86** | **[0.81-0.92]** | 0.85 | [0.69-1.06] | 1.12 | [0.75-1.66] |
| 80+ (vs 30-39) | **0.88** | **[0.86-0.90]** | **0.89** | **[0.82-0.97]** | **1.60** | **[1.34-1.90]** |
| Urban (vs rural) | **0.83** | **[0.81-0.85]** | **0.74** | **[0.70-0.78]** | 0.98 | [0.88-1.09] |
| Deprivation: 2 (vs 1) | **1.06** | **[1.04-1.08]** | 0.95 | [0.89-1.02] | **1.74** | **[1.45-2.10]** |
| 3 (vs 1) | **1.05** | **[1.03-1.07]** | **1.10** | **[1.02-1.20]** | **1.65** | **[1.38-1.97]** |
| 4 (vs 1) | **1.03** | **[1.01-1.05]** | **1.21** | **[1.12-1.32]** | **1.44** | **[1.21-1.71]** |
| 5 (vs 1) | **1.04** | **[1.02-1.07]** | **1.11** | **[1.02-1.20]** | 1.18 | [0.99-1.41] |
| 6 (vs 1) | **0.93** | **[0.90-0.95]** | **1.20** | **[1.10-1.31]** | **1.29** | **[1.08-1.55]** |
| 7 (vs 1) | 0.99 | [0.97-1.02] | **1.21** | **[1.11-1.31]** | 1.06 | [0.89-1.26] |
| 8 (vs 1) | 0.98 | [0.95-1.00] | **1.12** | **[1.03-1.23]** | 1.18 | [0.98-1.43] |
| 9 (vs 1) | **1.17** | **[1.14-1.20]** | 1.01 | [0.92-1.11] | 0.92 | [0.68-1.23] |
| 10 (vs 1) | **1.18** | **[1.14-1.22]** | 1.11 | [1.00-1.23] | 0.96 | [0.69-1.34] |

Notes: Ethnicity is excluded from the analysis due to large percentage of missing data. † The ‘other’ gender category is excluded for Scotland due to its small number.

Tables S8 Estimated odds ratios (ORs) and 95% Confidence intervals (CIs) from logistic regression models on intervention prescription (yes vs no) by country

|  | England  (n=141,011)  MI=30 | | Wales  (n=9,613)  MI=30 | | Scotland^†^  (n=2,370)  MI=30 | |
| --- | --- | --- | --- | --- | --- | --- |
|  | OR | 95% CI | OR | 95% CI | OR | 95% CI |
| Medical route (vs other) | **0.25** | **[0.24-0.26]** | **1.14** | **[1.00-1.31]** | **0.42** | **[0.32-0.56]** |
| Number of contacts | **1.07** | **[1.07-1.08]** | **1.17** | **[1.16-1.19]** | **1.19** | **[1.16-1.22]** |
| Mental health (vs no) | **0.97** | **[0.94-1.00]** | **0.30** | **[0.26-0.35]** | 0.85 | [0.70-1.03] |
| Physical health (vs no) | **1.05** | **[1.02-1.09]** | 0.94 | [0.83-1.06] | **1.41** | **[1.13-1.75]** |
| Social (vs no) | **0.93** | **[0.91-0.96]** | **0.64** | **[0.55-0.74]** | **0.60** | **[0.47-0.76]** |
| Lifestyle (vs no) | **0.93** | **[0.90-0.97]** | **0.54** | **[0.40-0.72]** | **2.03** | **[1.63-2.53]** |
| Employment (vs no) | **2.52** | **[2.41-2.63]** | **1.97** | **[1.41-2.74]** | 1.13 | [0.80-1.61] |
| Practical (vs no) | **0.88** | **[0.85-0.90]** | **2.47** | **[2.19-2.79]** | 0.86 | [0.66-1.12] |
| Other (vs no) | **0.95** | **[0.92-0.99]** | 1.13 | [0.96-1.32] | 0.81 | [0.61-1.08] |
| Male (vs female) | **0.96** | **[0.94-0.99]** | 0.92 | [0.83-1.02] | 1.03 | [0.85-1.25] |
| Other (vs female) | **1.75** | **[1.48-2.07]** | 1.23 | [0.55-2.75] | -- | -- |
| Age: 0-17 (vs 30-39) | **0.72** | **[0.66-0.80]** | **2.10** | **[1.65-2.67]** | **2.71** | **[1.07-6.85]** |
| 18-29 (vs 30-39) | **0.91** | **[0.87-0.95]** | **0.82** | **[0.68-0.97]** | 0.83 | [0.57-1.21] |
| 40-49 (vs 30-39) | **1.05** | **[1.01-1.09]** | 1.02 | [0.86-1.21] | 1.28 | [0.90-1.84] |
| 50-59 (vs 30-39) | **1.08** | **[1.04-1.13]** | 0.95 | [0.80-1.13] | 1.19 | [0.85-1.66] |
| 60-69 (vs 30-39) | **1.11** | **[1.07-1.16]** | 0.86 | [0.73-1.02] | 1.34 | [0.98-1.85] |
| 70-79 (vs 30-39) | 1.01 | [0.89-1.15] | **0.57** | **[0.33-1.00]** | 0.89 | [0.37-2.16] |
| 80+ (vs 30-39) | 0.92 | [0.87-0.96] | **0.47** | **[0.37-0.59]** | **0.63** | **[0.42-0.96]** |
| Urban (vs rural) | **1.27** | **[1.20-1.34]** | **0.76** | **[0.66-0.86]** | **0.73** | **[0.58-0.91]** |
| Deprivation: 2 (vs 1) | **0.53** | **[0.51-0.55]** | **1.53** | **[1.29-1.81]** | **0.53** | **[0.35-0.81]** |
| 3 (vs 1) | **0.47** | **[0.45-0.49]** | **1.31** | **[1.08-1.59]** | 0.70 | [0.47-1.03] |
| 4 (vs 1) | **0.46** | **[0.43-0.48]** | **1.53** | **[1.25-1.88]** | 1.10 | [0.76-1.59] |
| 5 (vs 1) | **0.49** | **[0.47-0.52]** | **1.55** | **[1.26-1.90]** | 0.94 | [0.65-1.37] |
| 6 (vs 1) | **0.41** | **[0.39-0.44]** | **1.90** | **[1.54-2.35]** | 0.84 | [0.57-1.23] |
| 7 (vs 1) | **0.49** | **[0.46-0.51]** | **1.52** | **[1.24-1.88]** | 1.29 | [0.89-1.88] |
| 8 (vs 1) | **0.57** | **[0.54-0.60]** | 1.14 | [0.91-1.42] | 0.95 | [0.63-1.44] |
| 9 (vs 1) | **0.49** | **[0.47-0.52]** | 1.27 | [1.01-1.61] | 0.71 | [0.37-1.35] |
| 10 (vs 1) | **0.54** | **[0.50-0.57]** | 1.03 | [0.78-1.34] | 0.77 | [0.37-1.61] |

Notes: Ethnicity is excluded from the analysis due to large percentage of missing data. † The ‘other’ gender category is excluded for Scotland due to its small number.

Table S9. Intervention costs in the UK and by country

| Cost | UK  (n=165,595) | England  (n=128,952) | Wales  (n=9,227) | Scotland  (n=4,263) | Northern Ireland  (n=10,474) |
| --- | --- | --- | --- | --- | --- |
| Free | 91.7% | 95.6% | 95.9% | 50.3% | 78.7% |
| Out-of-pocket cost | 6.3% | 3.4% | 3.2% | 21.8% | 18.6% |
| Subsidised | 1.9% | 1.1% | 1.0% | 28.0% | 2.8% |

Figure S1. Sample selection diagram

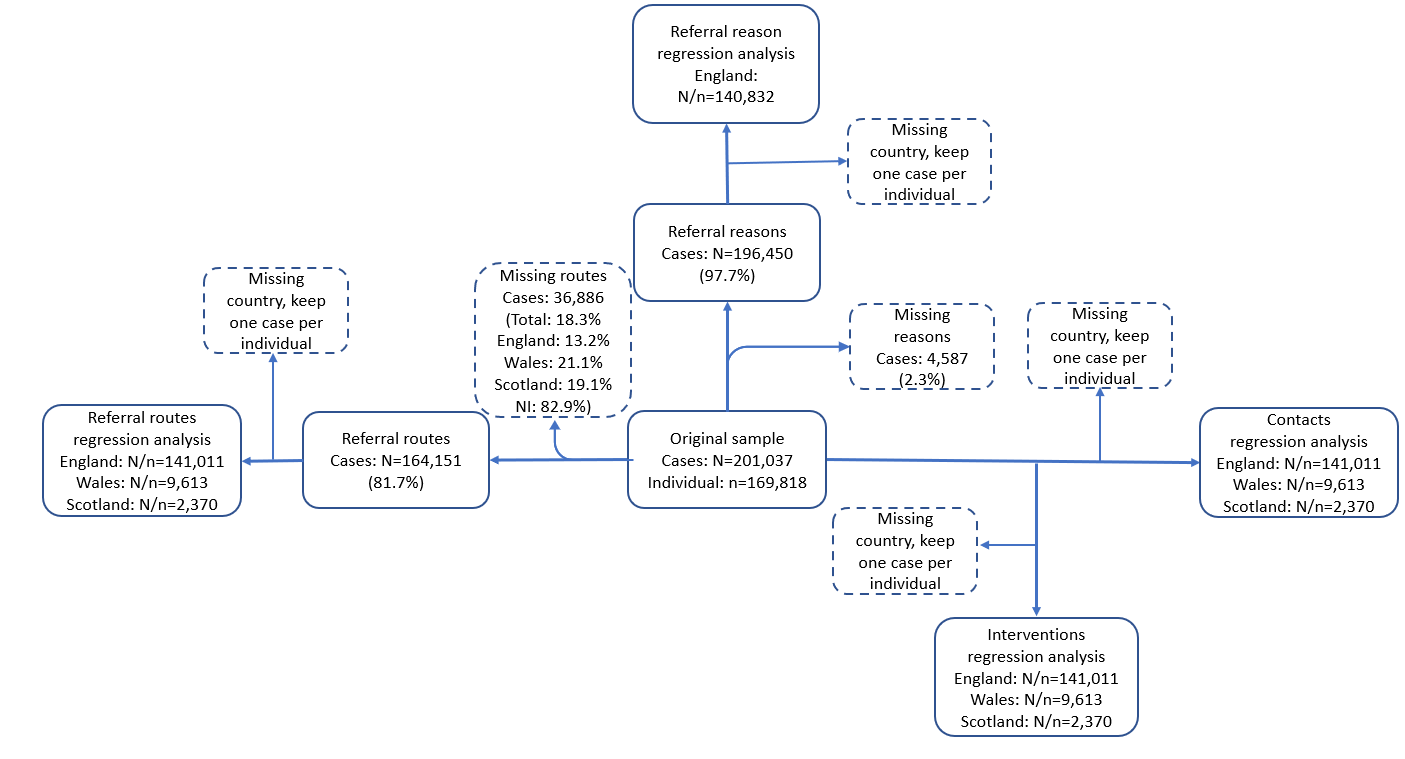

Figure S2. Number of clients in each local authority across the UK (N=160,145)

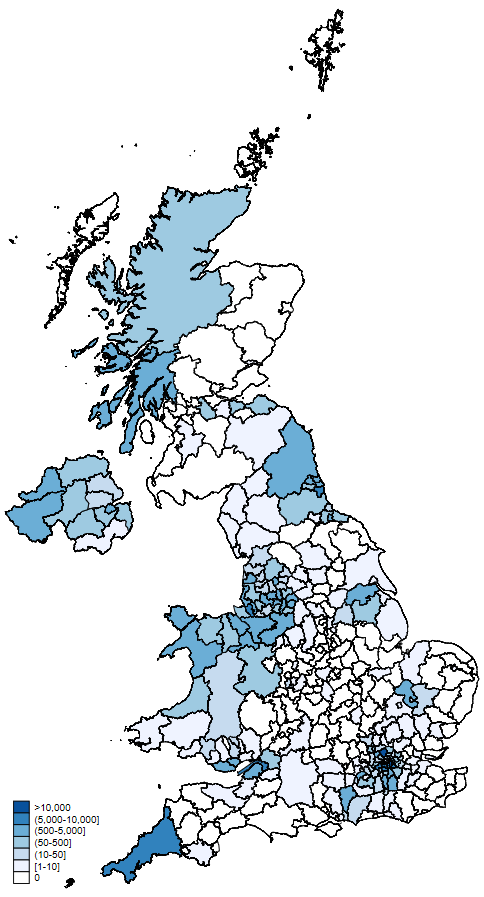

Notes: *Based upon Land Cover Map 2019 © UKCEH 2020. Contains Ordnance Survey data © Crown Copyright 2007, License number 100017572.*

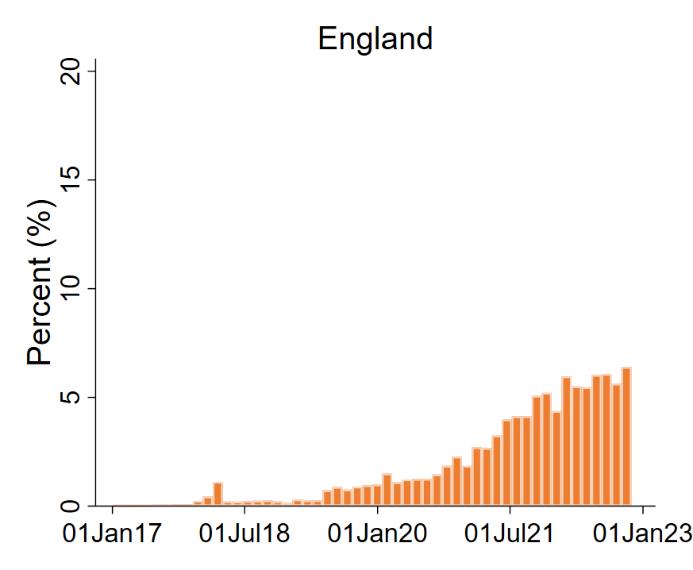

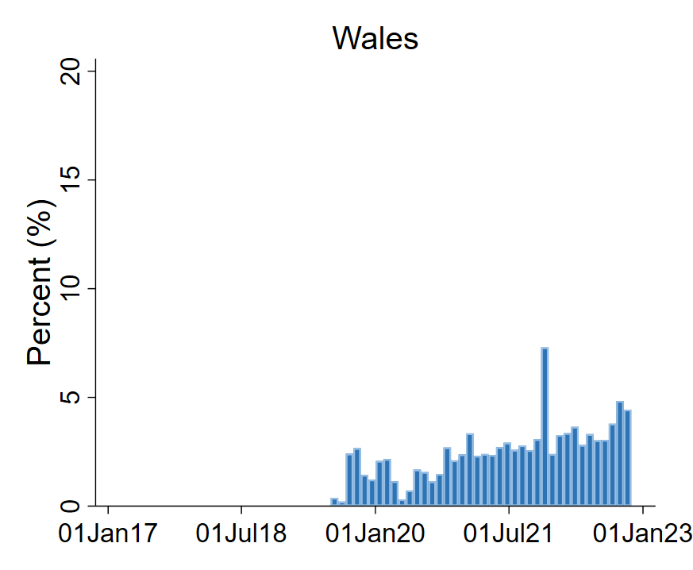

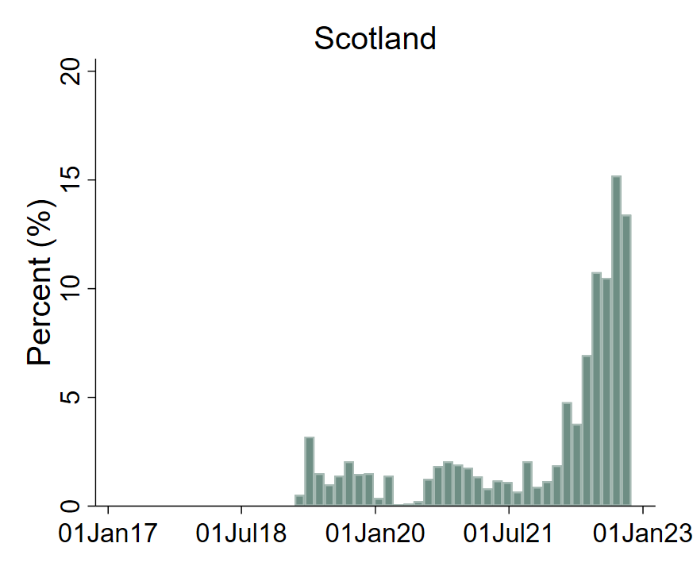

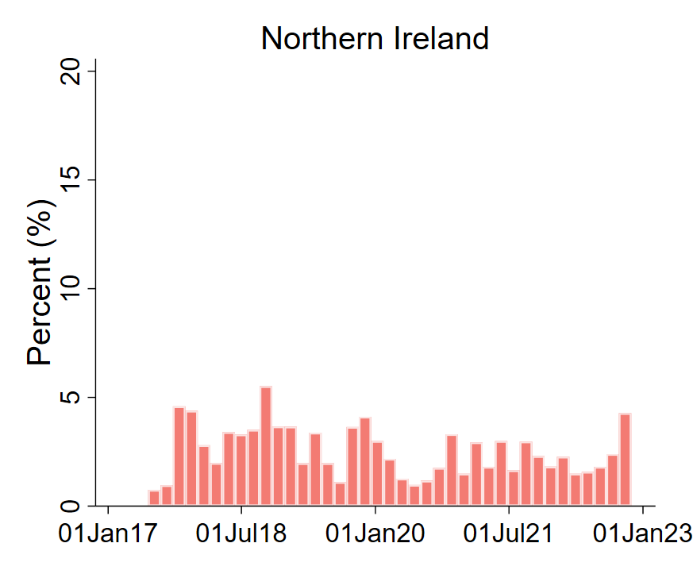

Figure S3. Date distribution of social prescribing cases in the UK and by countries January 2017-Novermber 2022

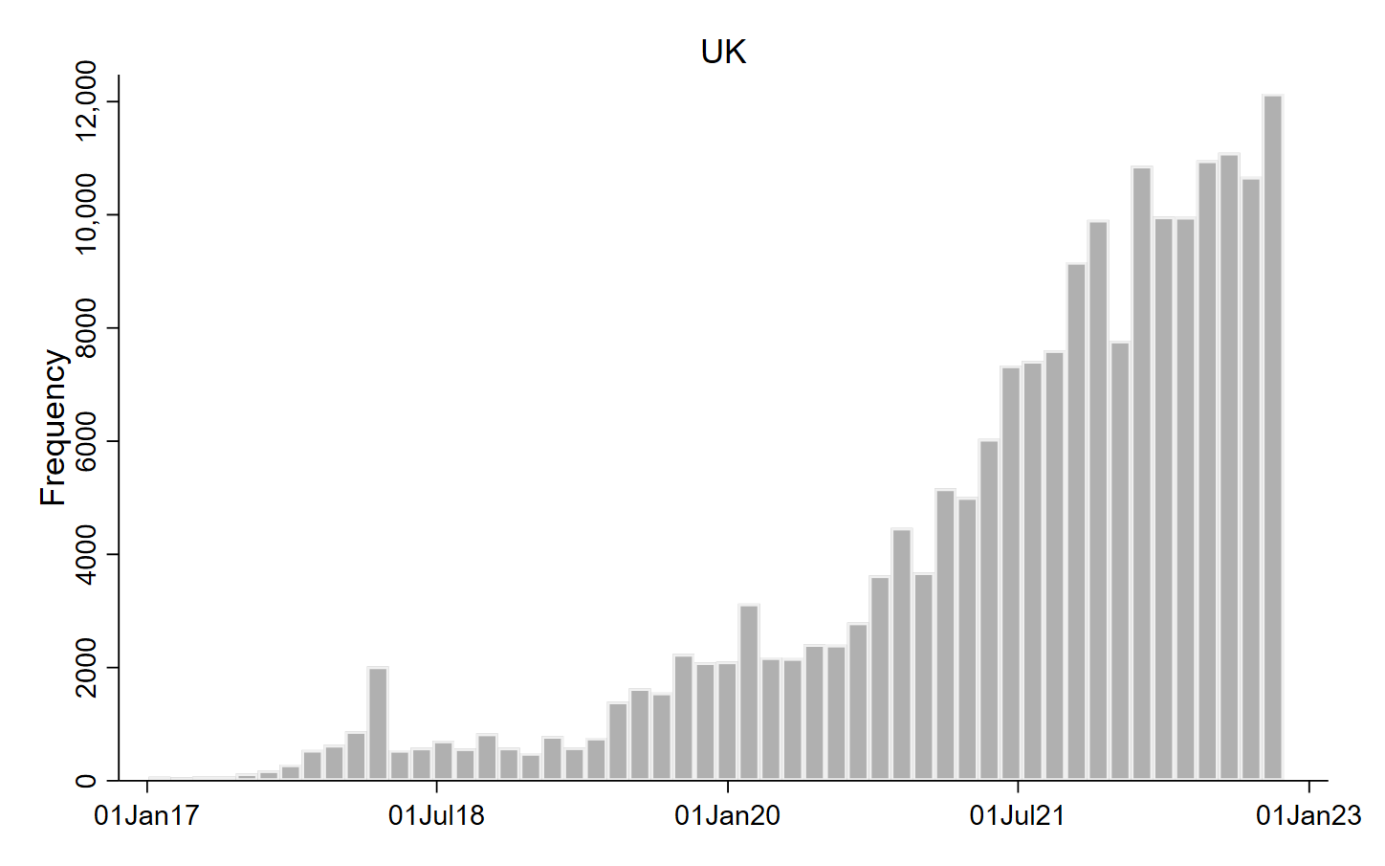

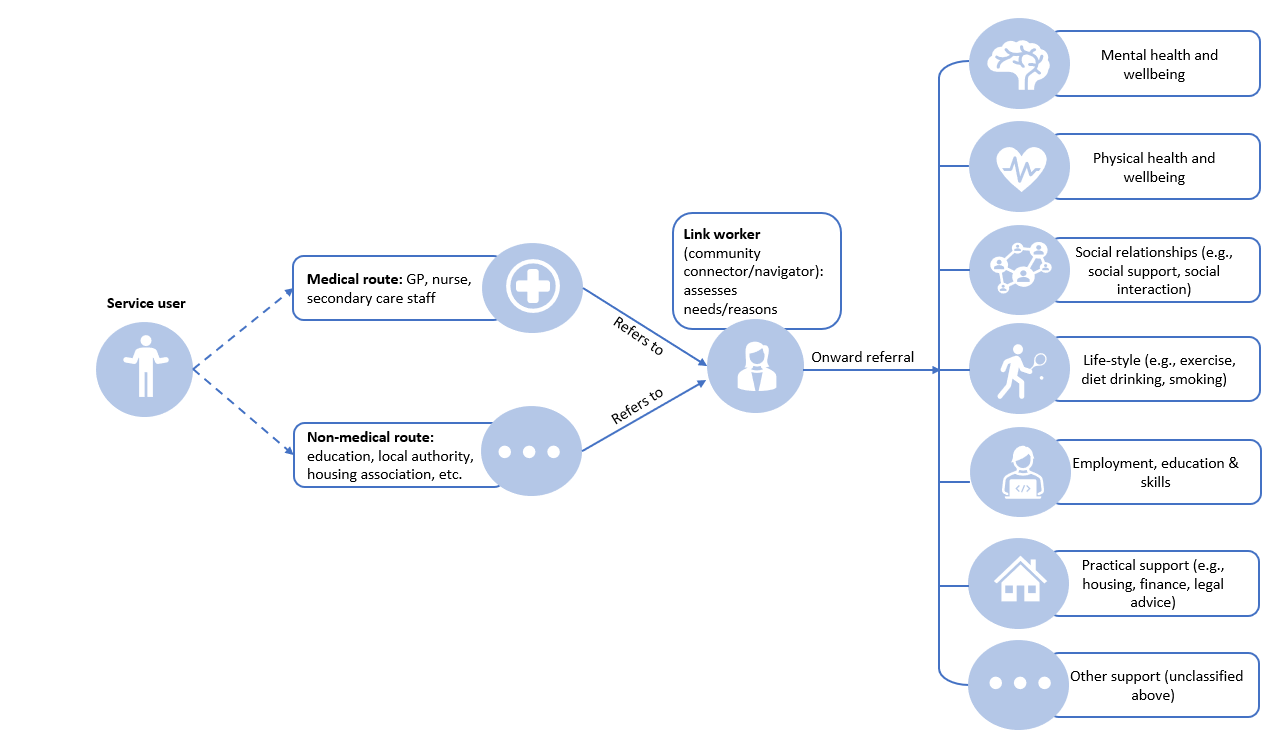

Figure S4. Social prescribing medical and non-medical pathways in the UK

Figure S5 Percentage of cases with referral reasons from each domain by country

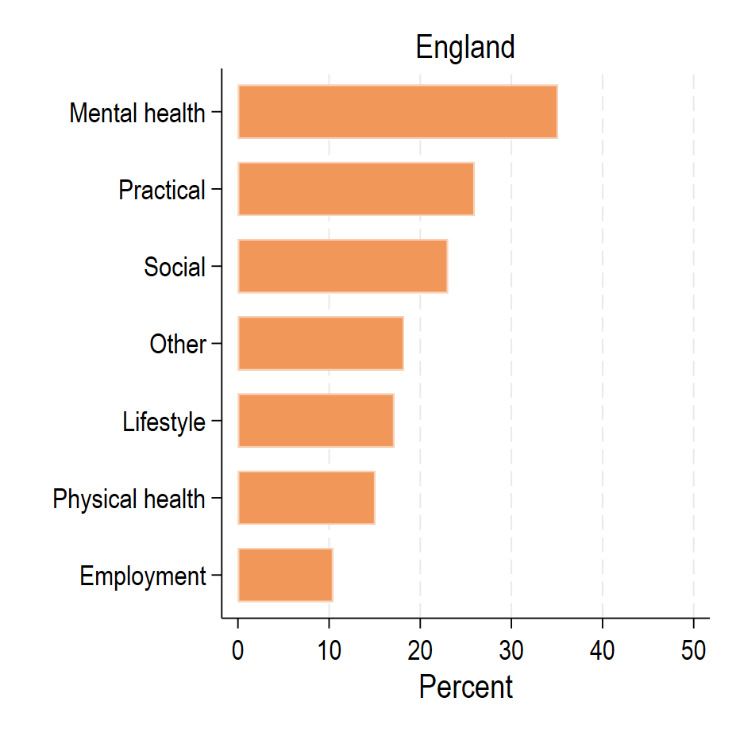

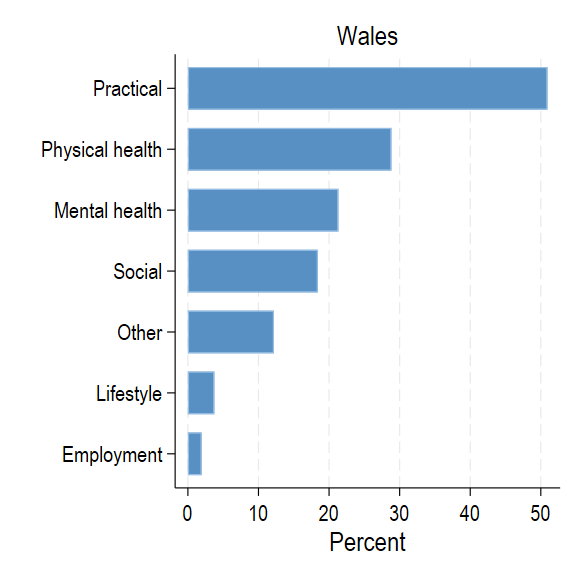

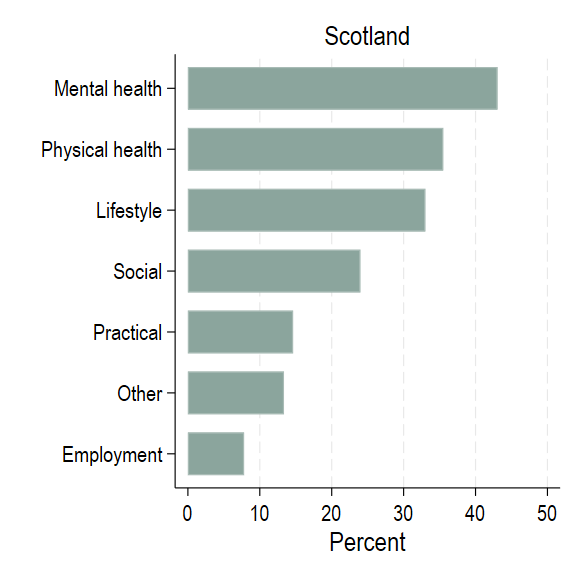

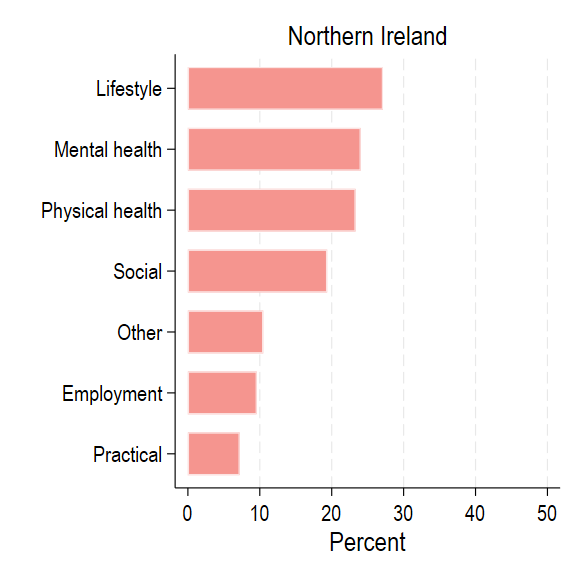

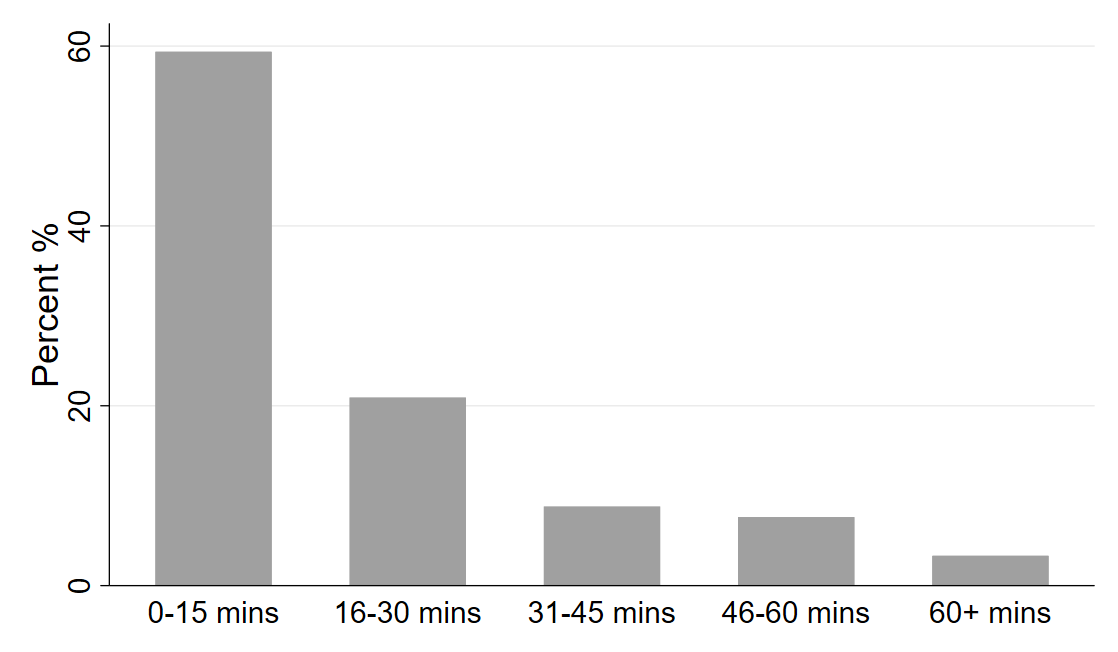

Figure S6. Distribution of contact time with link workers

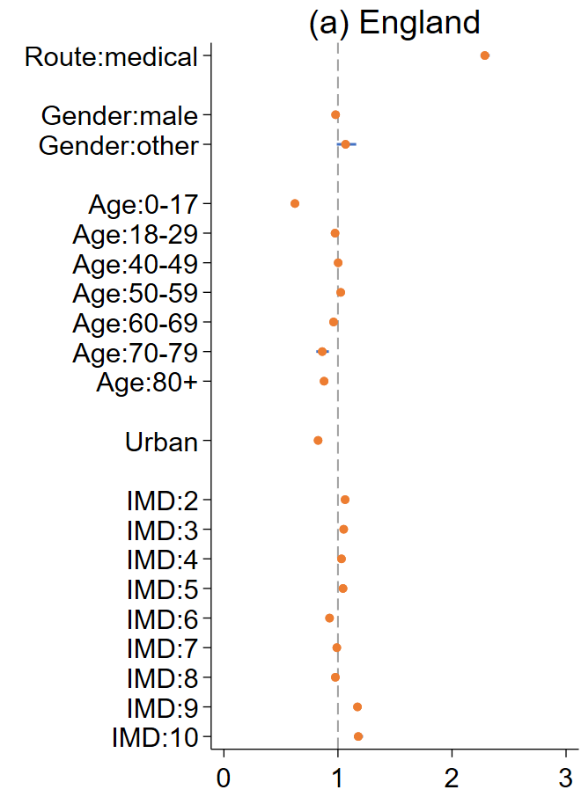

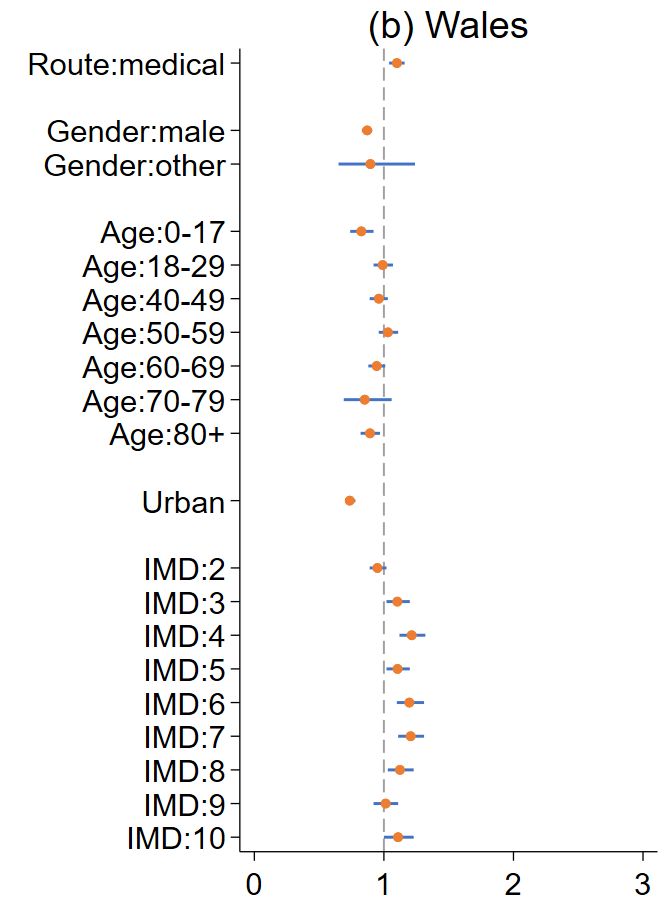

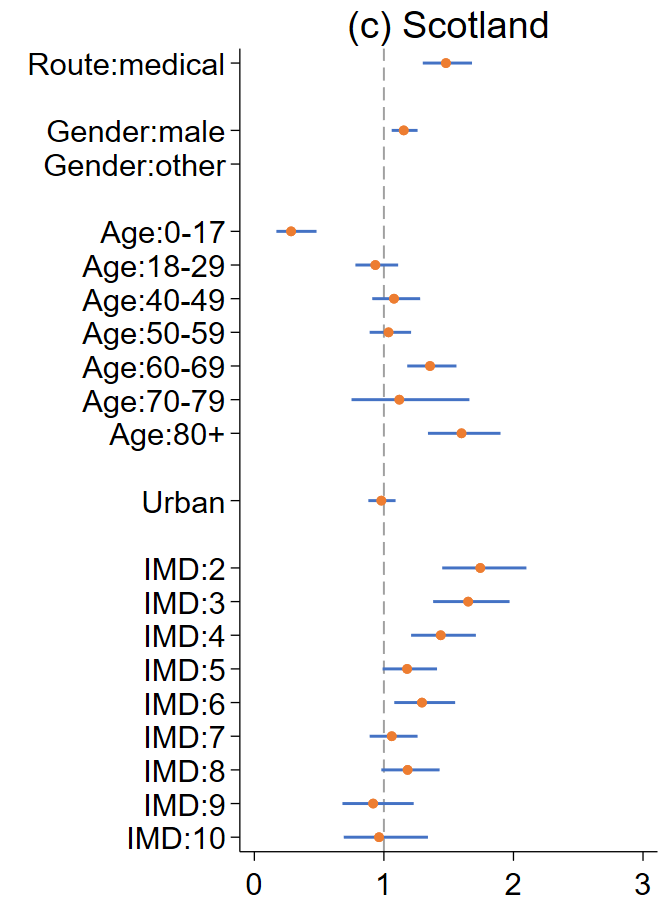

Figure S7 Incidence rate ratios and 95% confidence intervals from the negative binomial regression model on the number of contacts with link workers by country

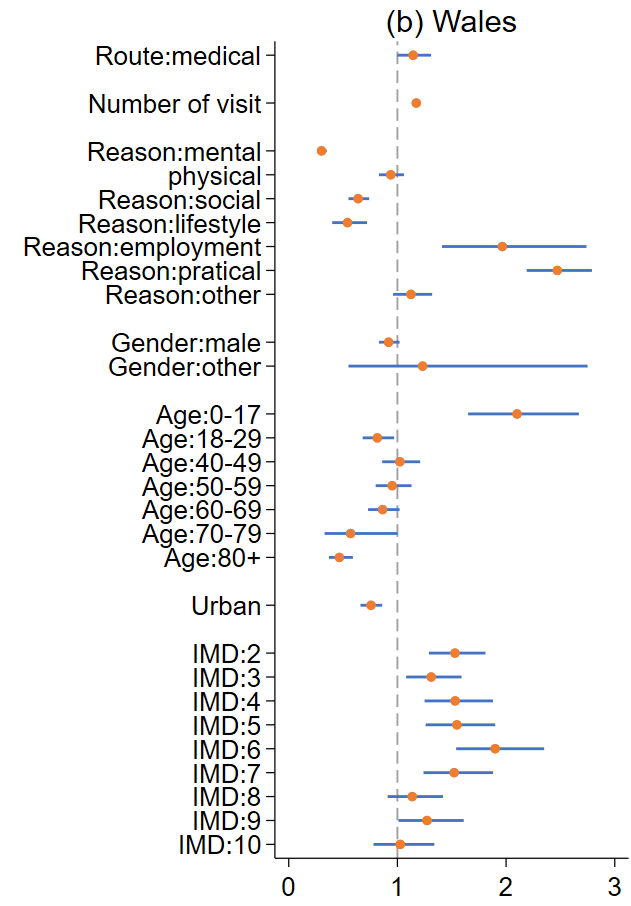

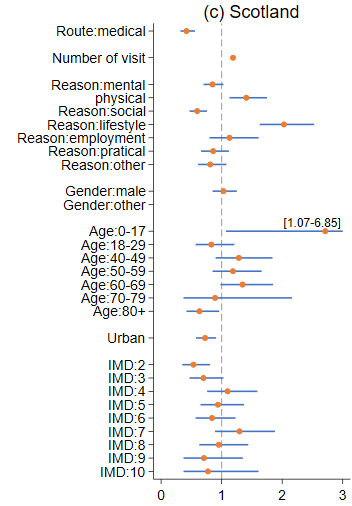

Figure S8 Odds ratios and 95% confidence intervals from the logistic regression model on having an intervention prescribed by country

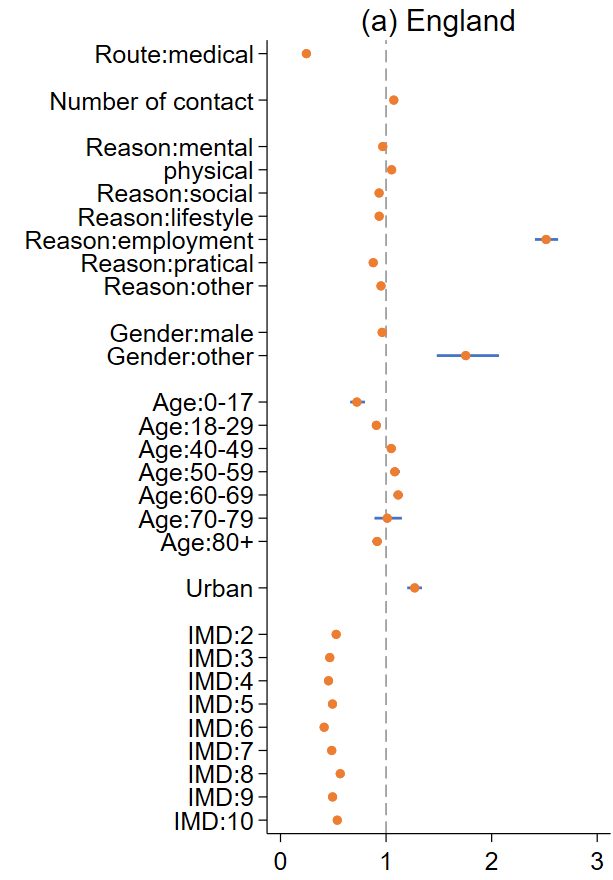

Figure S9. Percentages of intervention related to referral reasons by domain by country

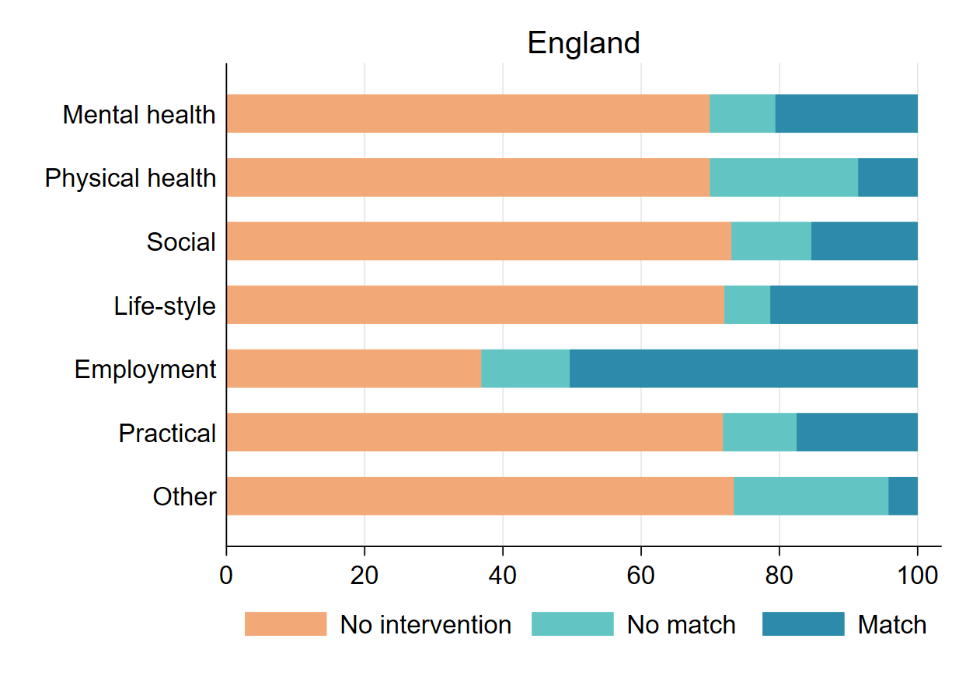

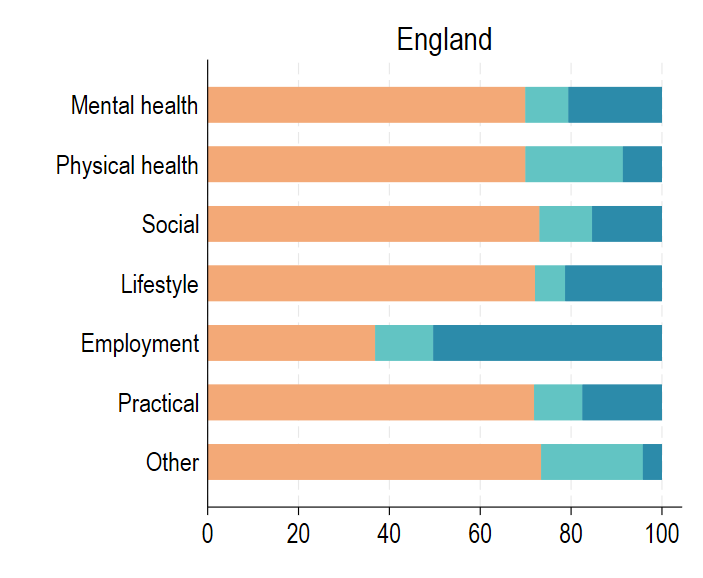

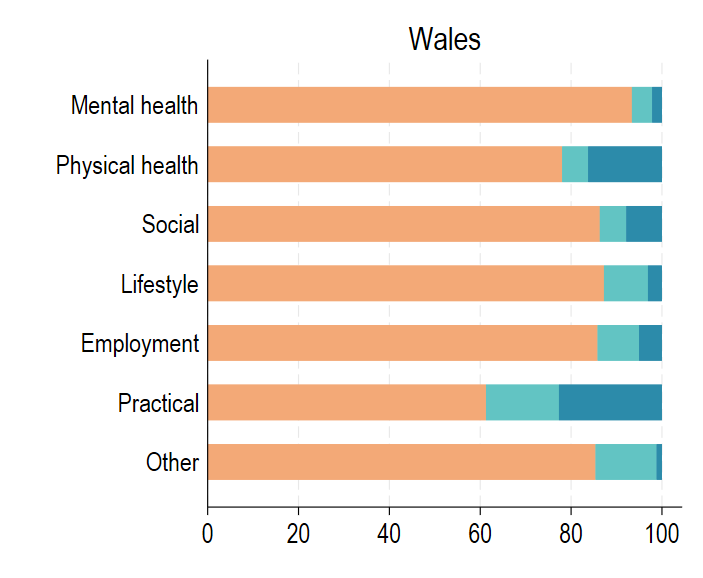

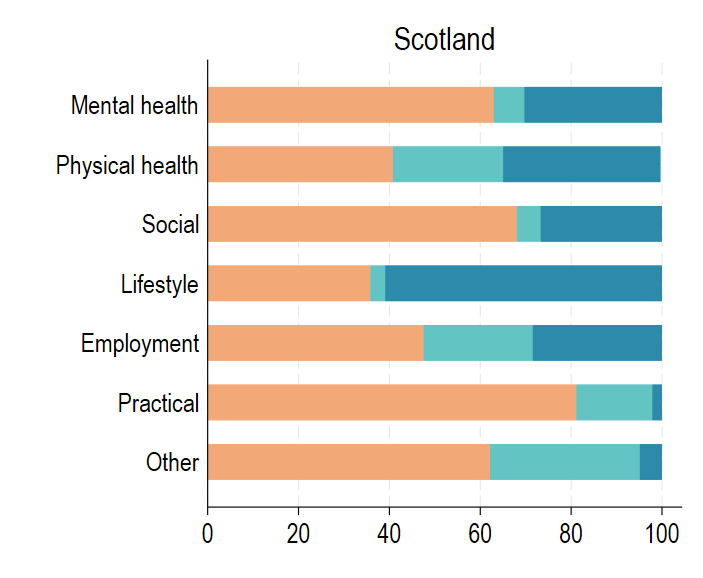

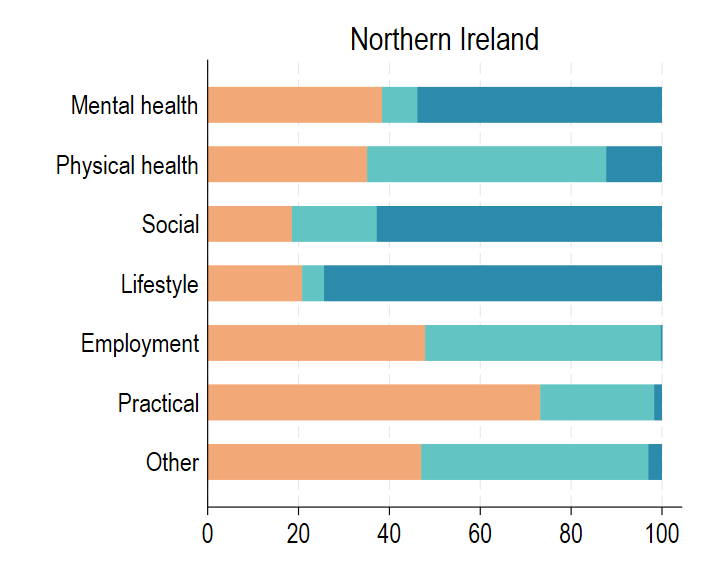
